# Beyond the TyG Index: Composite Metabolic Metrics Integrating Central Adiposity Improve Atrial Fibrillation Risk Prediction Independent of Genetic Susceptibility

**DOI:** 10.64898/2026.04.02.26350088

**Authors:** Zhang Ke, Shengwei Wang, Wang Song, Shifeng Zhao, Meng He, Changwei Ren, Hao Cui, Yongqiang Lai

**Affiliations:** Department of Cardiovascular Surgery, Beijing Anzhen Hospital, Capital Medical University; Beijing Anzhen Hospital, Capital Medical University, The Key Laboratory of Remodeling-Related Cardiovascular Diseases, Ministry of Education, Beijing Institute of Heart, Lung and Blood Vessel Diseases, Beijing 100029, China

**Keywords:** Atrial fibrillation, Insulin resistance, TyG index, Obesity, Polygenic risk score

## Abstract

**Background:** Insulin resistance (IR) and obesity are key drivers of atrial fibrillation (AF). However, the comparative predictive value of the Triglyceride-Glucose (TyG) index versus composite indices combining IR and anthropometric measures—such as TyG-BMI, TyG-Waist Circumference (TyG-WC), and Waist-to-Height Ratio (WHtR)—remains undefined. We aimed to evaluate these associations and the modifying effect of genetic susceptibility.

**Methods:** We analyzed 293,318 UK Biobank participants free of AF at baseline. Hazard ratios (HRs) were estimated using Cox proportional hazards models, and non-linearity was assessed using restricted cubic splines. Incremental predictive value was evaluated via Net Reclassification Improvement (NRI). Interactions with AF Polygenic Risk Scores (PRS) were examined.

**Results:** During follow-up, 22,707 incident AF cases occurred. While the TyG index was associated with AF in unadjusted models, this association was nullified after full adjustment. In contrast, composite indices (TyG-BMI, TyG-WC) and WHtR showed robust, positive associations (WHtR HR per SD: 1.30, 95% CI 1.28–1.32). Spline analysis identified non-linear threshold effects (e.g., WHtR inflection at 0.556). Adding WHtR or TyG-BMI to baseline models significantly improved risk reclassification (NRI ∼10.3–11.8%, P<0.001), whereas TyG alone did not (P=0.73). Elevated metabolic risk increased AF incidence across all genetic categories, with significant interactions suggesting greater relative impact in low-genetic risk groups.

**Conclusions:** Composite indices integrating central obesity and insulin resistance are superior to the TyG index alone in predicting incident AF. The identification of specific risk thresholds and genetic interactions highlights "metabolic health" as a crucial, modifiable target for AF prevention.

**Research Insights:** 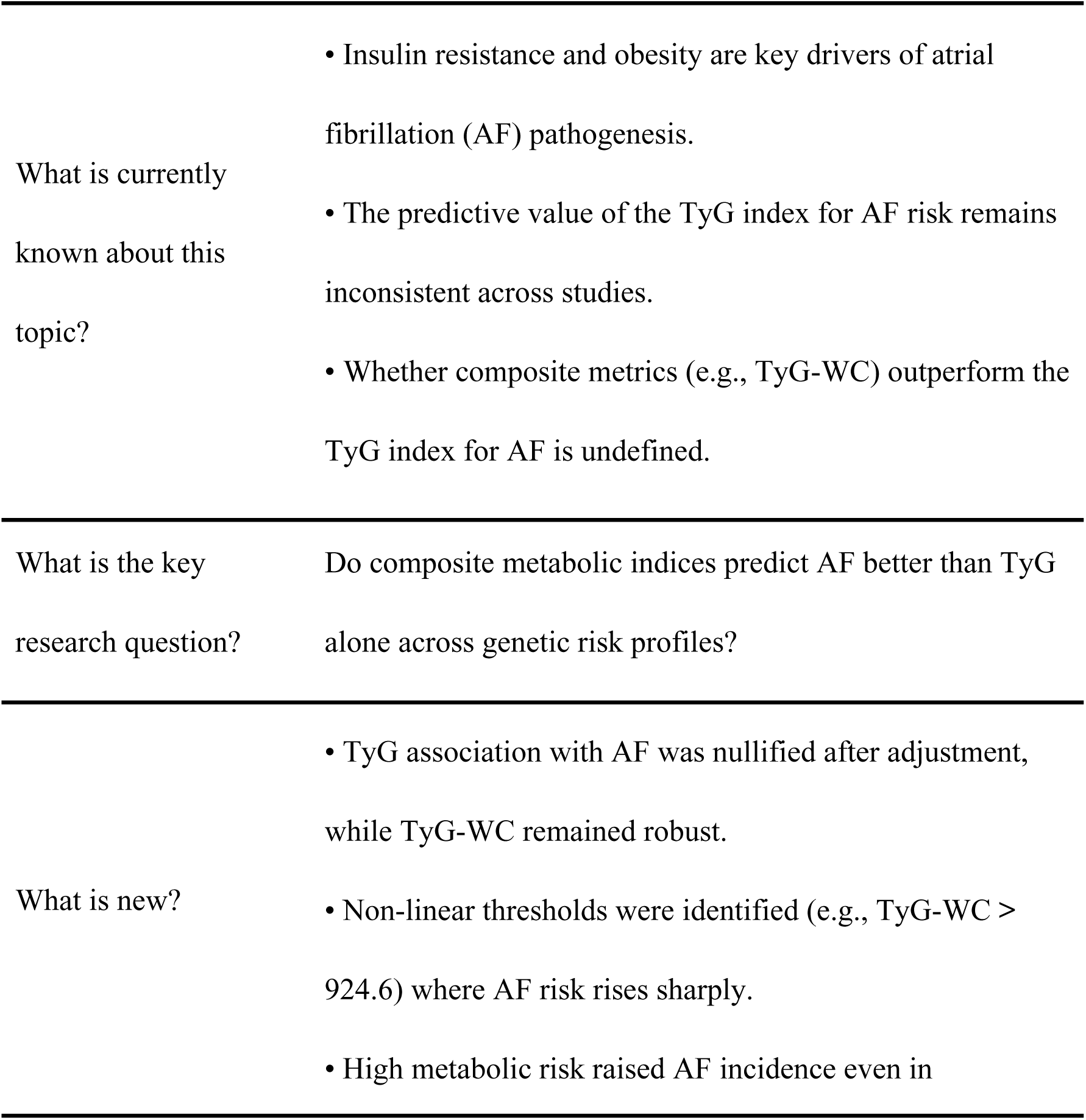

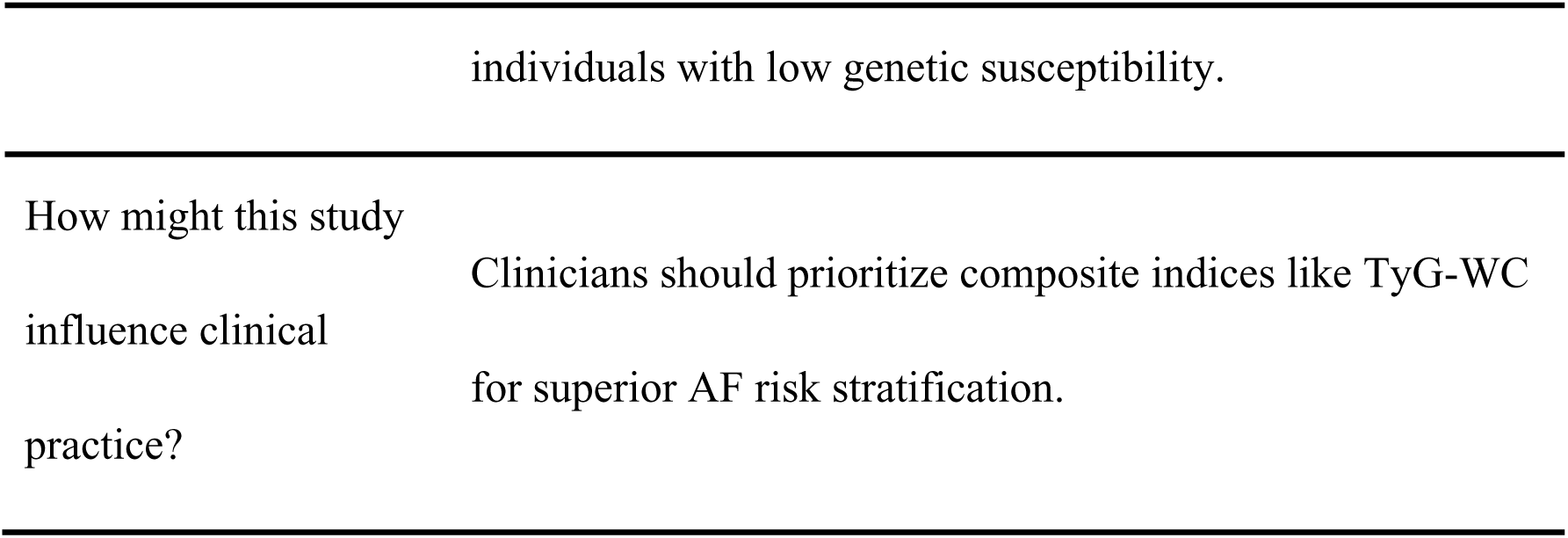

## 1. Introduction

Atrial fibrillation (AF) is a major global health burden and a leading cause of stroke and heart failure^1,2^. While traditional risk factors explain a portion of AF liability, metabolic dysregulation—specifically insulin resistance (IR) and adiposity—has emerged as a central driver of arrhythmogenesis^3,4^. This link is reinforced by the cardiovascular-kidney-metabolic (CKM) syndrome framework and the elevated AF risk observed in severe insulin-resistant diabetes^5,6^. Mechanistically, this is driven by systemic inflammation, oxidative stress, and adipose-induced atrial remodeling.

Given the pivotal role of IR, the triglyceride-glucose (TyG) index has gained traction as an accessible surrogate marker^7^. While meta-analyses link elevated TyG to AF risk, evidence regarding its independent predictive value remains inconsistent^8^. A key limitation of the TyG index is that it captures hepatic and peripheral IR but overlooks the critical role of adipose tissue distribution. Visceral and epicardial adipose tissues are not merely energy stores but active endocrine organs that secrete pro-inflammatory adipokines and promote atrial fibrosis^4,9^.

To address this limitation, composite indices integrating IR with anthropometry—such as TyG-BMI, TyG-Waist Circumference (TyG-WC), the cardiometabolic index (CMI), and the metabolic score for visceral fat (METS-VF)—were developed to capture the synergistic "dual-hit" of IR and adiposity^10,11^. However, a systematic head-to-head comparison of these composite indices against the standalone TyG index in large-scale populations is lacking.

Furthermore, the dose-response relationships and potential non-linear risk thresholds remain undefined^12,13^. Additionally, the interaction between these modifiable metabolic risks and fixed genetic susceptibility, quantified by polygenic risk scores (PRS), represents a critical knowledge gap. It remains unclear whether metabolic indices contribute to risk through pathways distinct from genetic variants^14^.

Using the UK Biobank resource, we conducted a comprehensive evaluation of seven single and composite metabolic indices for predicting incident AF. We assessed their discriminatory power, reclassification improvement, and non-linear thresholds, while also examining the modifying effect of genetic susceptibility. This study aims to determine if combining anthropometric and IR measures provides a more robust framework for AF risk stratification.

## 2. Methods

### 2.1 Study Population

The current study leveraged data derived from the UK Biobank, a large-scale prospective cohort consisting of more than 500,000 participants recruited from assessment centres throughout England, Scotland, and Wales (2006–2010). Data acquisition involved touchscreen-based questionnaires, physical examinations, and the collection of biological specimens. Detailed cohort characteristics have been described elsewhere^15^. The study protocol received ethical approval from the North West Multicentre Research Ethics Committee, and all participants gave written informed consent before enrollment. This analysis was carried out under UK Biobank Application Number 545415.

Starting with an initial cohort of 501,936 subjects, we subsequently excluded individuals with prevalent AF (n=8,461). Further exclusions were made for participants lacking complete data on IR-related indices (n=73,798), covariates (n=68,349), or PRS (n=58,010). Consequently, the final study population comprised 293,318 participants (Figure 1).

**Figure 1.**
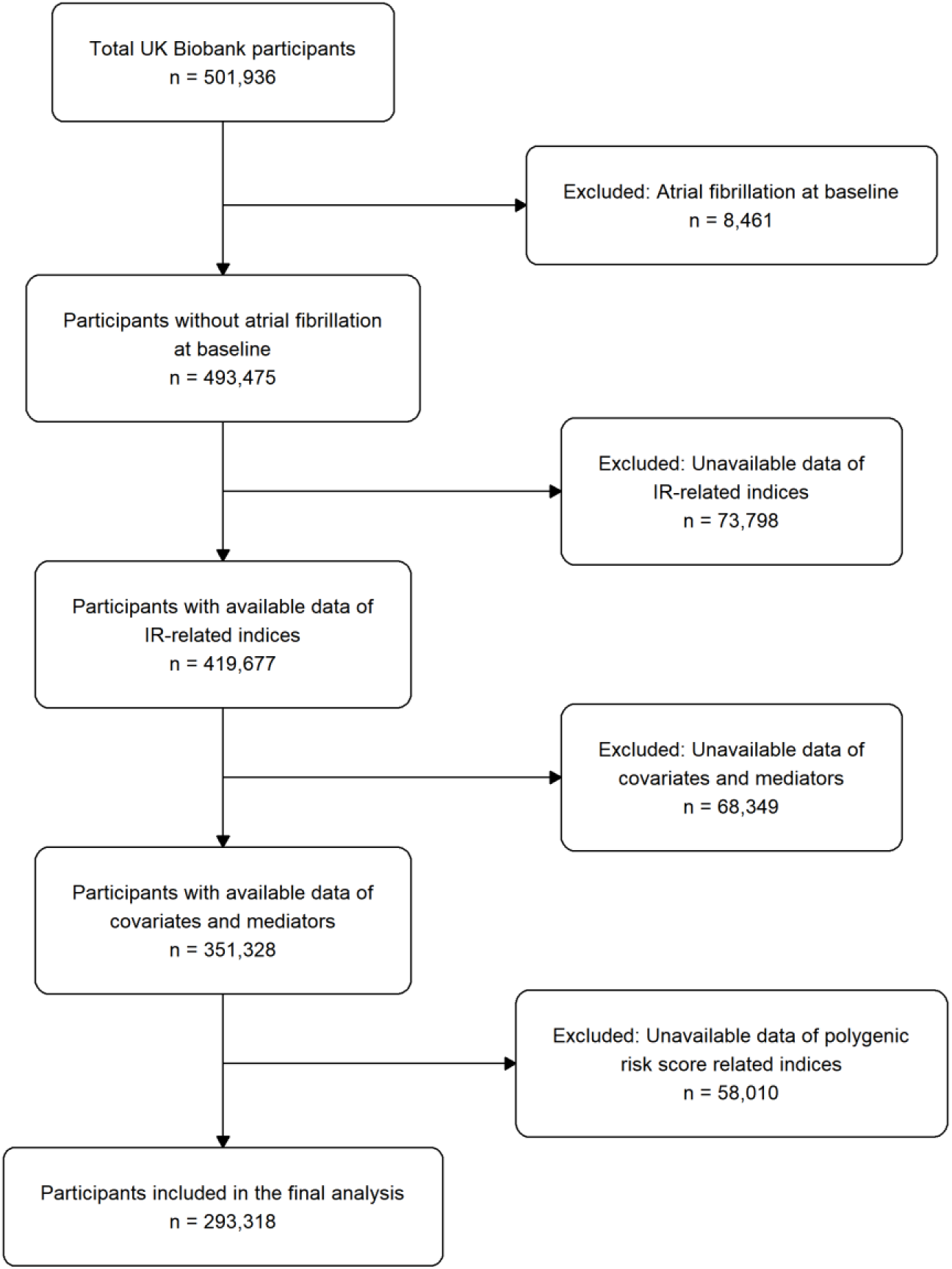
Study design and criteria. The diagram depicts the step-by-step recruitment process. From the initial cohort of 501,936 participants, individuals were sequentially excluded if they had prevalent atrial fibrillation at baseline, missing data on insulin resistance-related indices (TyG, BMI, WC, etc.) or covariates, or lacked genetic data for Polygenic Risk Score (PRS) calculation. The final analytic cohort consisted of 293,318 participants. Abbreviations: AF, atrial fibrillation; PRS, polygenic risk score; TyG, triglyceride-glucose index.

### 2.2 Exposure Assessment

The following indices were calculated:

We assessed multiple surrogates of insulin resistance. The TyG index was determined using the formula: ln [fasting triglyceride (mg/dL) * fasting glucose (mg/dL) / 2], while the Waist-to-Height Ratio (WHTR) was calculated by dividing waist circumference by height. Additional indices, including TyG-BMI, TyG-WC, CMI, and Metabolic Score for Insulin Resistance (METS-IR), were derived using standard equations referenced in prior studies^16–19^. Regarding genetic susceptibility, the PRS was divided into tertiles to classify participants into low, intermediate, or high risk categories.

### 2.3 Outcome Ascertainment

We defined the primary outcome as incident AF or atrial flutter (ICD-10 code I48). Cases were ascertained via linkage to hospital inpatient records and national death registries. Follow-up duration was calculated from the date of recruitment to the earliest of the following endpoints: diagnosis of AF/atrial flutter, death, loss to follow-up, or the end of the study period (September 25, 2024).

### 2.4 Statistical Analysis

Baseline demographic and clinical characteristics were summarized using descriptive statistics, stratified by incident AF status. Continuous variables are presented as mean ± standard deviation (SD), while categorical variables are expressed as counts (percentages). Differences between groups were evaluated using Student’s t-tests for normally distributed continuous data, Mann-Whitney U tests for non-normally distributed data, and Pearson’s chi-square tests for categorical variables.

We utilized univariate and multivariable Cox proportional hazards models to evaluate the association between IR-related indices and the risk of incident AF. Exposures were analyzed both as quartiles and as continuous variables standardized per SD increment. Three models were constructed: Model 1 (unadjusted), Model 2 (adjusted for age and sex), and Model 3 (further adjusted for Townsend Deprivation Index (TDI), smoking status, and alcohol consumption). Kaplan-Meier (KM) curves were plotted to visualize the cumulative incidence of AF across quartiles, and differences between groups were assessed using the log-rank test. We characterized potential non-linear dose-response relationships using restricted cubic spline (RCS) functions with four knots, overlaying density histograms to visualize participant distribution. In instances where non-linearity was detected, a two-piece Cox regression model was utilized to determine threshold inflection points. To enhance clinical interpretability, hazard ratios were rescaled to relevant unit increments (e.g., per 0.1 unit for WHTR; per 10 units for TyG-BMI and METS-IR; and per 100 units for TyG-WC). We further evaluated the predictive accuracy of the metabolic indices via time-dependent receiver operating characteristic (ROC) curve analyses, calculating the area under the curve (AUC) for 3-, 5-, and 10-year AF risk. Finally, the incremental predictive value was assessed using Net Reclassification Improvement (NRI) and Integrated Discrimination Improvement (IDI).

We utilized the Standard PRS for AF provided by the UK Biobank, which was constructed using a Bayesian approach (PRS-CS) based on the largest available GWAS summary statistics. To minimize population stratification bias, the analysis was strictly restricted to participants of genetically defined Caucasian ancestry. To facilitate interpretation and ensure sufficient statistical power within subgroups for interaction analyses, participants were categorized into tertiles based on the PRS distribution of the entire cohort: Low risk (lowest tertile), Intermediate risk (middle tertile), and High risk (highest tertile), with the lowest tertile serving as the reference group for subsequent interaction analyses. To account for residual population structure, the first ten genetic principal components were extracted and standardized (mean of 0, standard deviation of 1) prior to their inclusion as adjustment covariates in the multivariable models.

Subgroup and Sensitivity Analyses We conducted stratified analyses to evaluate the consistency of associations across key demographic and lifestyle subgroups, including age (< 65 vs. ≥65 years), smoking status, and alcohol consumption (categorized as never, previous, or current). To verify the robustness of our results and mitigate potential reverse causality, we performed sensitivity analyses by excluding participants diagnosed with AF within the first two years of follow-up. Further sensitivity assessments applying Fine-Gray subdistribution hazard models to account for death as a competing risk.

All statistical computations were performed using R software, version 4.5.2 (R Foundation for Statistical Computing). For biomarker-related analyses, statistical significance was defined by a False Discovery Rate (FDR)-adjusted P-value < 0.05 (Benjamini-Hochberg procedure). For all other analyses, a two-sided P-value < 0.05 was considered statistically significant.

## 3. Results

### 3.1 Baseline Characteristics

A total of 293,318 participants were included in the final analysis, among whom 22,707 (7.7%) developed incident AF during the follow-up period. The baseline characteristics of the study population, stratified by AF status, are presented in Table 1. Compared with the non-AF group, participants who developed AF were significantly older (mean age 61.9 vs. 56.4 years) and had a higher proportion of males (62.0% vs. 44.8%). The AF group exhibited a more adverse cardiovascular risk profile, characterized by higher systolic and diastolic blood pressure, higher Townsend deprivation index, and a greater prevalence of previous smoking compared to controls (all P < 0.001). Notably, regarding the primary exposures of interest, the incident AF group demonstrated significantly elevated levels across all seven surrogate markers of insulin resistance and obesity. Specifically, TyG, TyG-BMI, TyG-WC, CMI, WHTR, TyG-WHTR, and METS-IR were all significantly higher in participants with incident AF compared to those without (all P < 0.001). Furthermore, participants in the AF group had higher levels of HbA1c, C-reactive protein (CRP), and white blood cell counts, but lower levels of LDL-C and platelet counts compared with the control group (all P < 0.001).

**Table 1.**
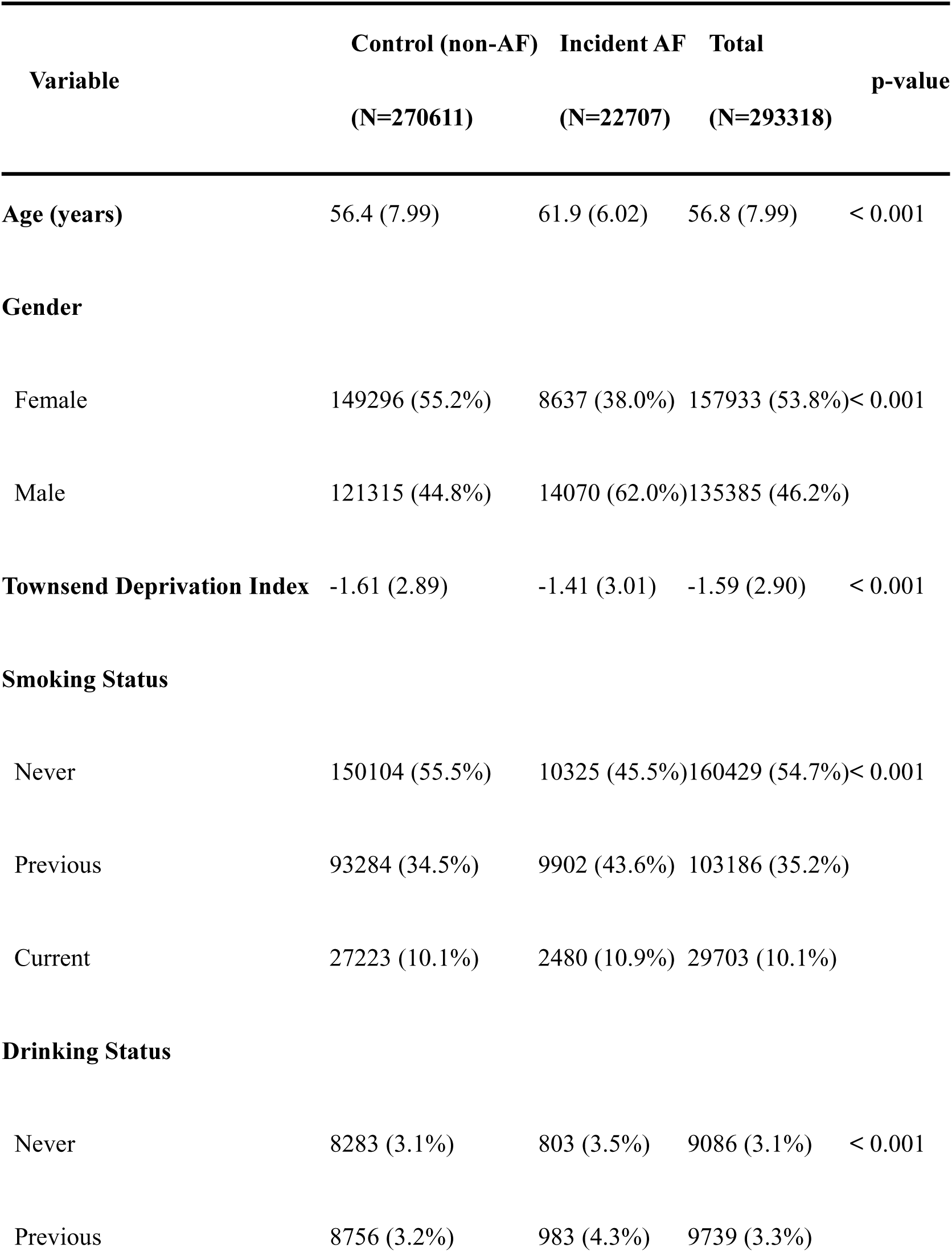
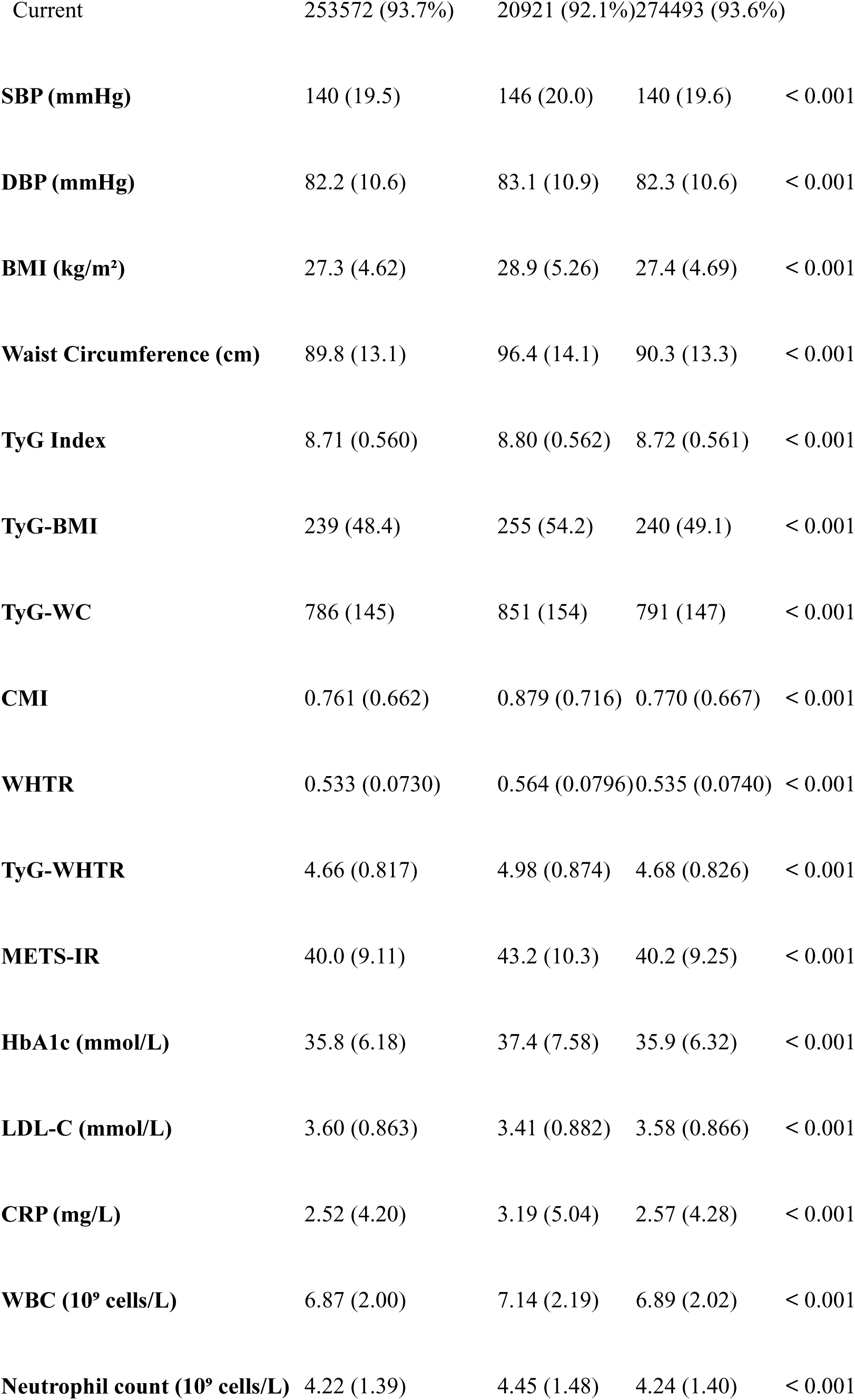
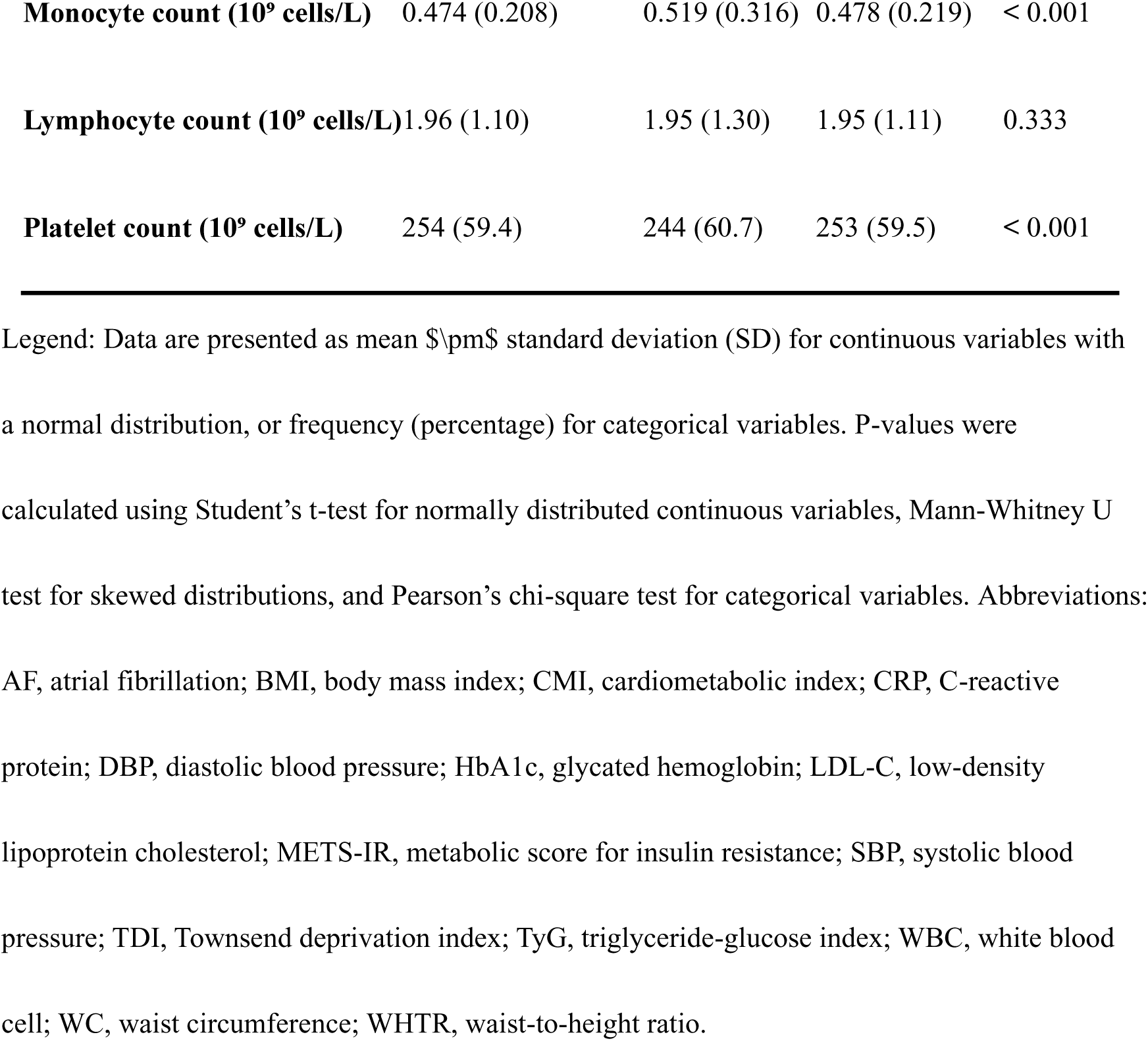
Baseline characteristics of the study population stratified by incident atrial fibrillation status.

### 3.2 Cumulative Incidence of AF

Kaplan-Meier survival analyses were performed to visualize the cumulative incidence of AF across quartiles of the seven metabolic and anthropometric indices (Figure 2). For all investigated indices—TyG, WHTR, TyG-BMI, TyG-WHTR, CMI, TyG-WC, and METS-IR—participants in the highest quartile (Q4) exhibited a significantly higher cumulative risk of incident AF compared to those in the lowest quartile (Q1) (all log-rank tests P < 0.001). The divergence in AF incidence curves among the quartile groups became apparent early in the follow-up period and progressively widened over time, suggesting a robust stratification ability of these markers for AF risk. Detailed numbers of at-risk participants at each follow-up time point across quartiles of the indices are provided in Supplementary Table S1.

**Fig 2.**
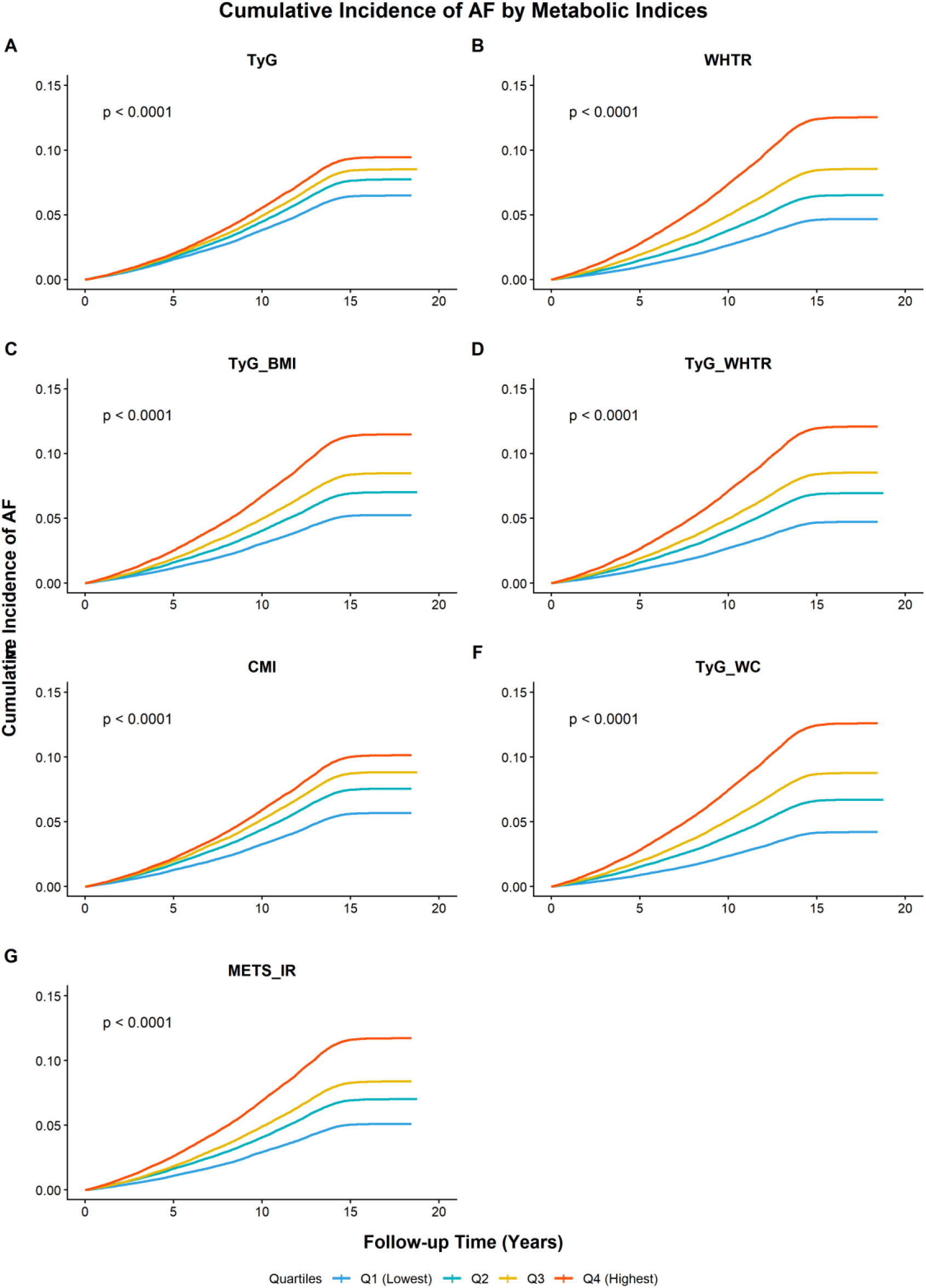
Kaplan-Meier curve of surrogate markers of insulin resistance and obesity for AF risk. Cumulative incidence estimates of new-onset atrial fibrillation (AF) stratified by quartiles (Q1–Q4) of seven metabolic indices: (A) TyG index, (B) Waist-to-Height Ratio (WHtR), (C) TyG-BMI, (D) TyG-WHtR, (E) Cardiometabolic Index (CMI), (F) TyG-WC, and (G) METS-IR.

**Figure 3.**
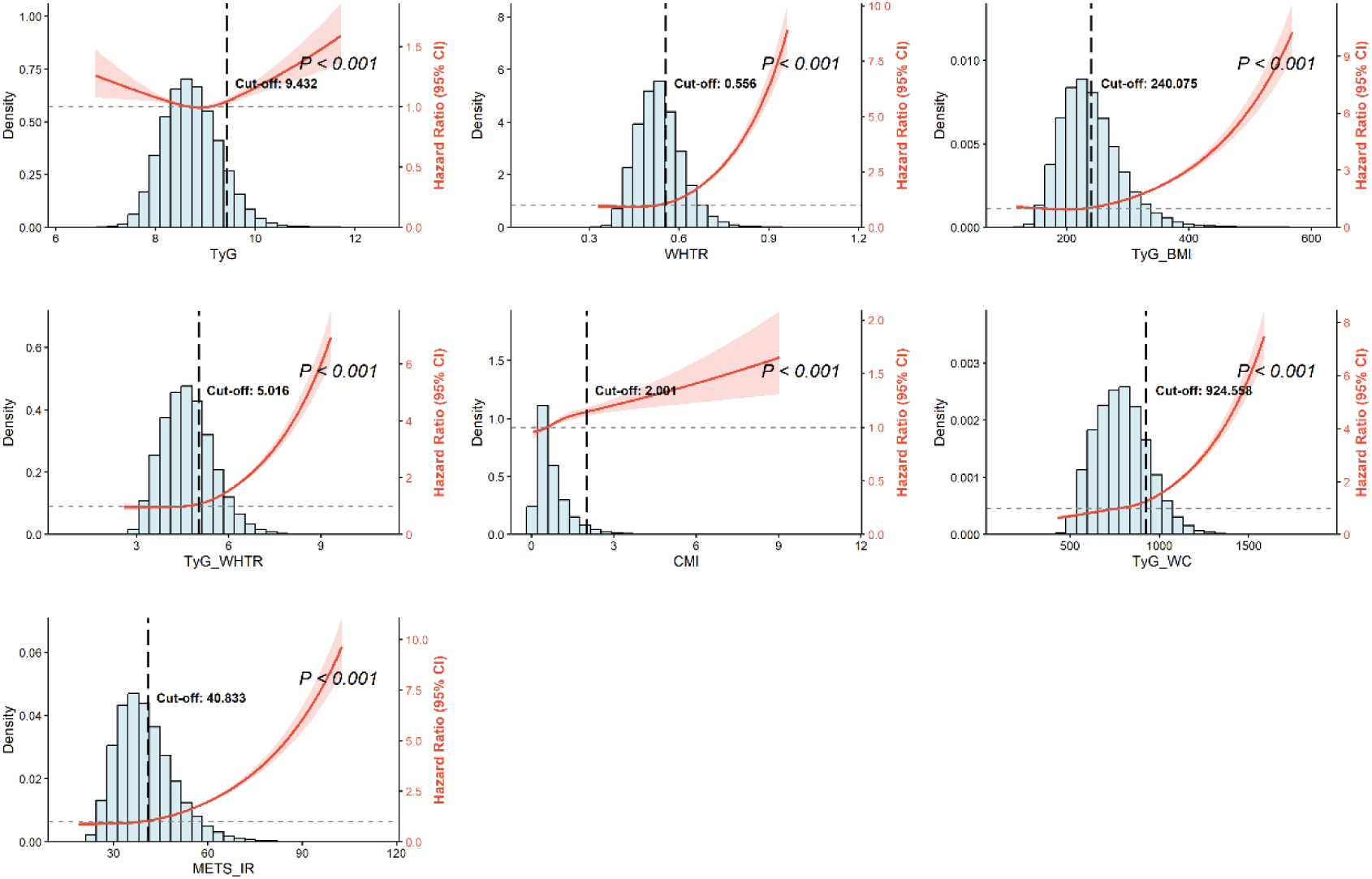
RCS curve with dose-response plot between IR-related indices and risk of AF occurrence. Restricted cubic spline (RCS) plots visualizing the hazard ratios (HRs, solid red lines) and 95% confidence intervals (shaded red areas) for incident AF across the continuous distribution of metabolic indices. Histograms (blue bars) show the distribution of participants. Models were fully adjusted for age, sex, deprivation index, lifestyle factors, and comorbidities (Model 3). Dashed vertical lines mark the identified inflection points (cut-off values) where the risk trajectory significantly changes. Note: The TyG index shows a J-shaped association, whereas composite indices (e.g., TyG-WC, TyG-BMI) demonstrate a monotonic but non-linear increase, with risk accelerating sharply beyond the identified thresholds. P-values indicate the significance of the non-linear term.

The x-axis represents the follow-up duration in years, and the y-axis represents the cumulative incidence rate. For all indices, participants in the highest quartile (Q4, red line) exhibited a significantly higher risk of AF compared to those in the lowest quartile (Q1, blue line), with the divergence widening over time. P-values were calculated using the log-rank test. Abbreviations: BMI, body mass index; WC, waist circumference; METS-IR, metabolic score for insulin resistance.

### 3.3 Association between Indices and AF

The associations between the seven metabolic indices and the risk of incident AF were evaluated using Cox proportional hazards models (Table 2). In the fully adjusted models (Model 3), which controlled for potential confounders including age, sex, socioeconomic status, lifestyle factors, and comorbidities, all investigated indices were significantly and positively associated with the risk of developing AF. When analyzed as continuous variables, each 1-SD increase in TyG-WC and METS-IR showed the strongest associations with AF risk (HR [95% CI]: [1.32 (1.30-1.34)] and [1.30 (1.28-1.31)], respectively; all P < 0.001). Consistently, this positive correlation persisted when the indices were categorized into quartiles. Participants in the highest quartile (Q4) of all seven indices had a significantly increased risk of AF compared to those in the lowest quartile (Q1). The trend analysis demonstrated a robust dose-response relationship (P for trend < 0.001for all indices), suggesting that higher cumulative burdens of insulin resistance and central obesity are independent predictors of AF onset.

**Table 2.**
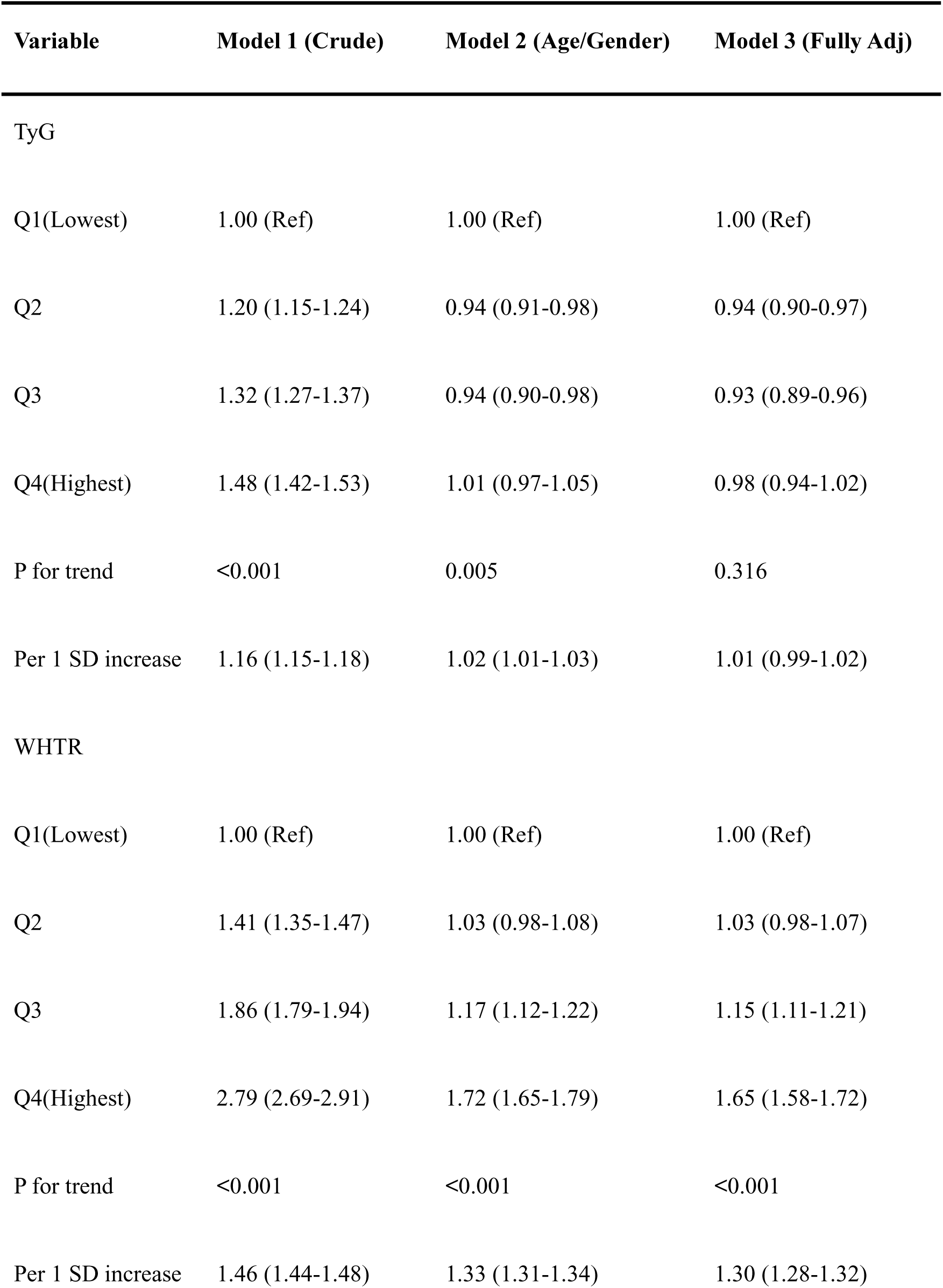
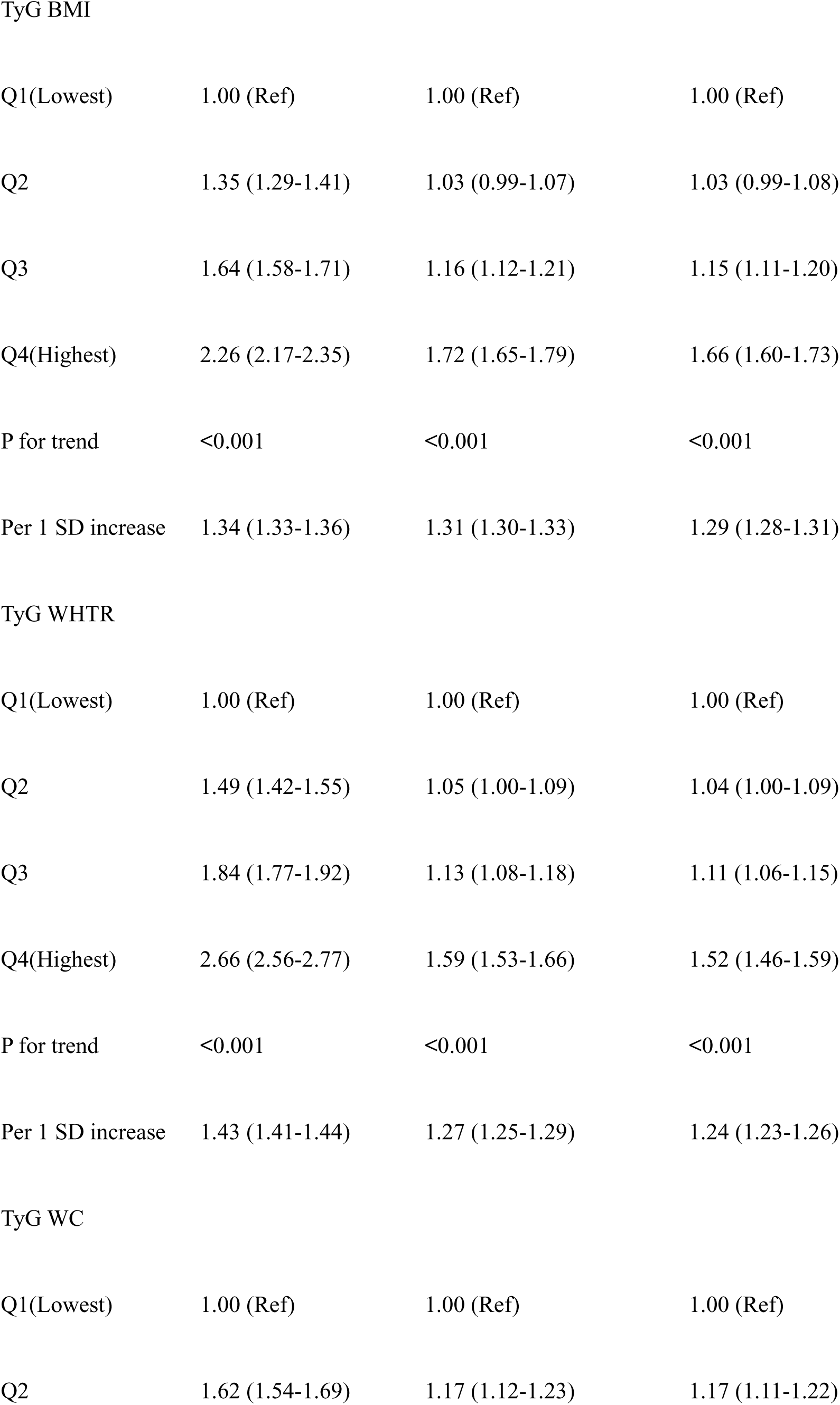
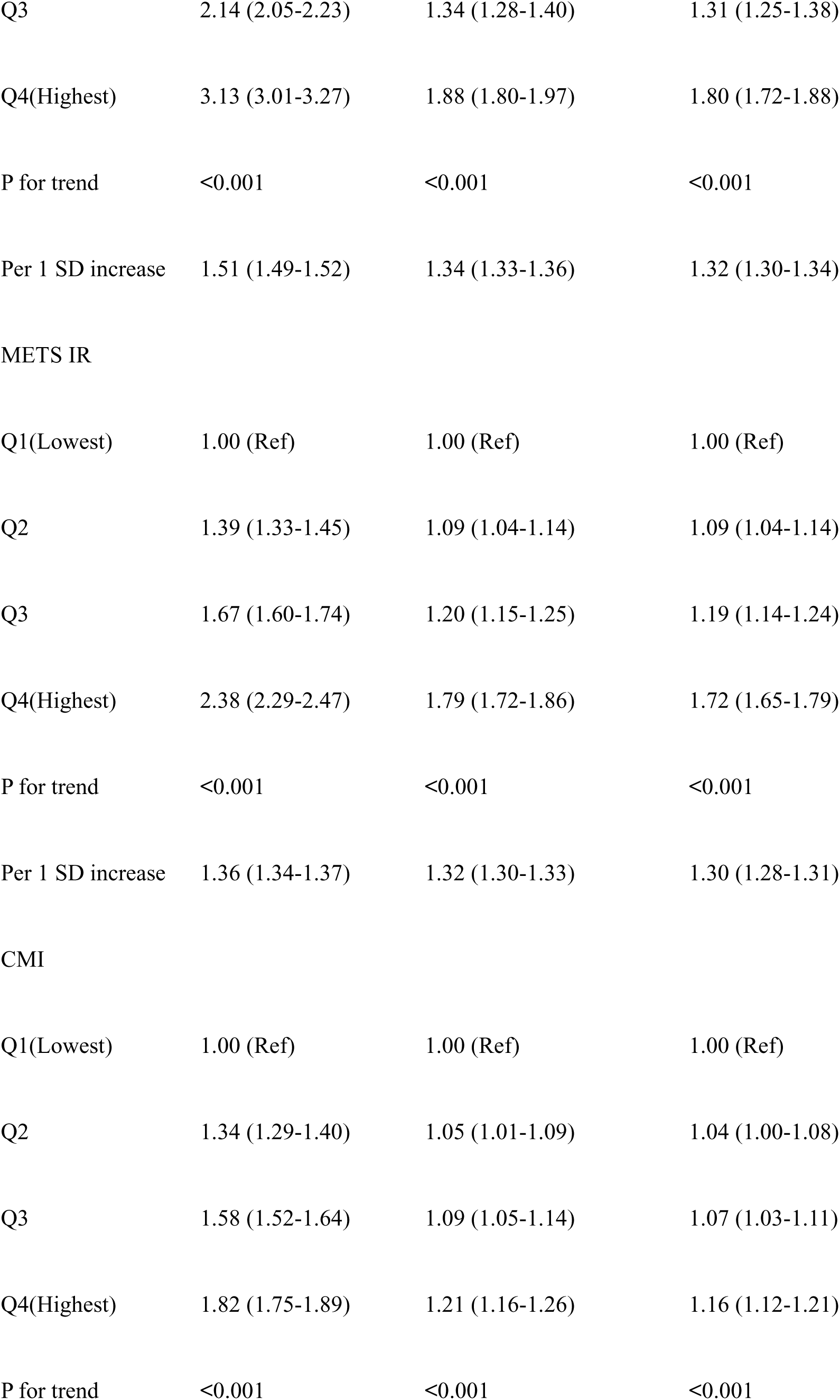
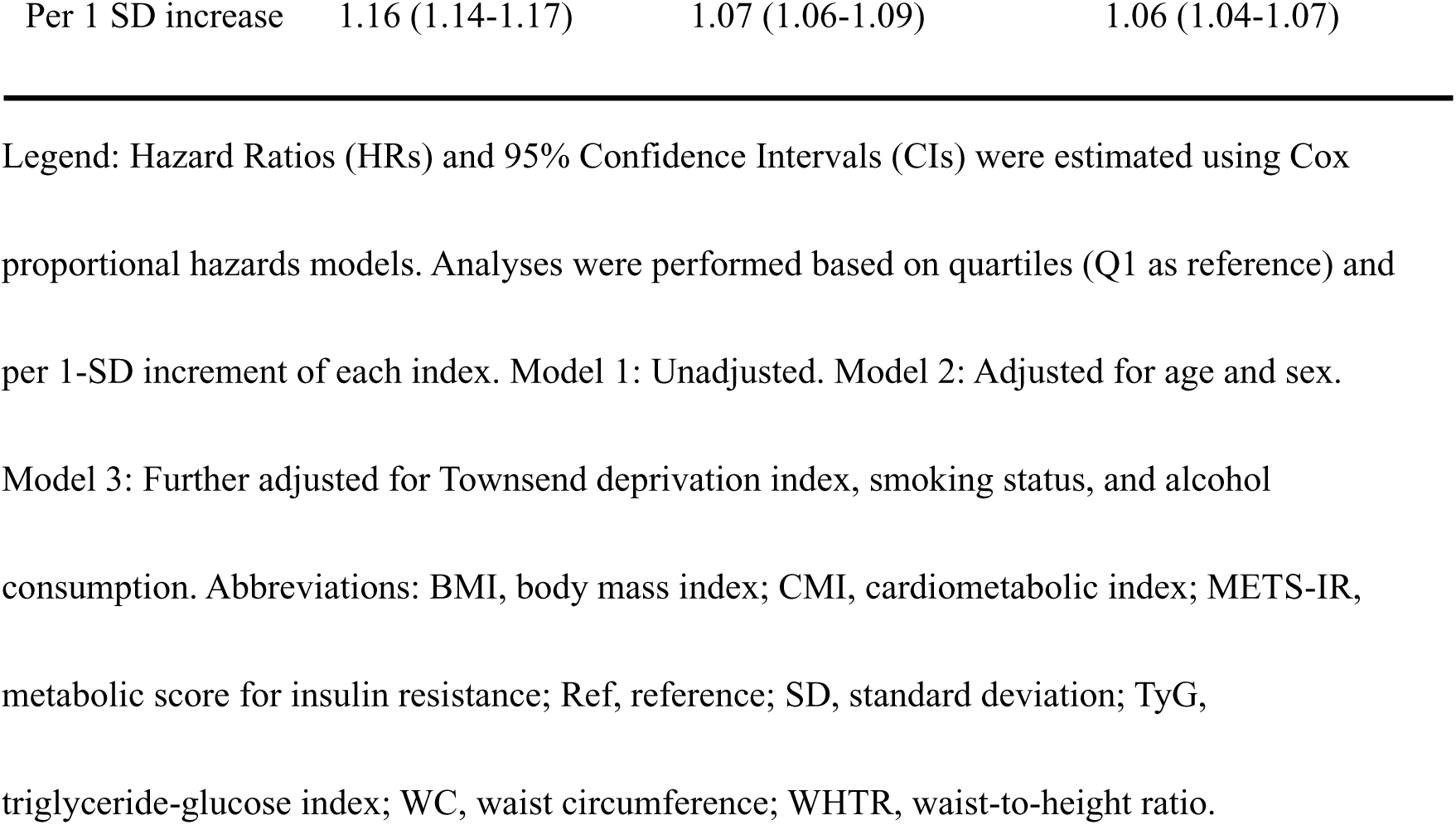
Associations between insulin resistance-related metabolic indices and risk of incident atrial fibrillation.

### 3.4 Non-linear Relationships and Threshold Analysis

To further characterize the nature of these associations, we utilized restricted cubic splines (RCS) based on the fully adjusted models (Model 3). As illustrated in Figure 2, significant non-linear relationships were identified for all investigated indices (all P for non-linearity < 0.001). The TyG index exhibited a distinctive J-shaped association with incident AF. Threshold analysis identified an inflection point at 9.432, shown in Table 3. Below this threshold, a higher TyG index was associated with a decreased risk of AF (HR: 0.954, 95% CI: 0.926–0.983, P = 0.002). Conversely, above the inflection point, the risk of AF increased significantly with elevated TyG levels (HR: 1.341, 95% CI: 1.231–1.462, P < 0.001), indicating that strictly controlling TyG levels within a specific range is crucial for AF prevention.

**Table 3.**
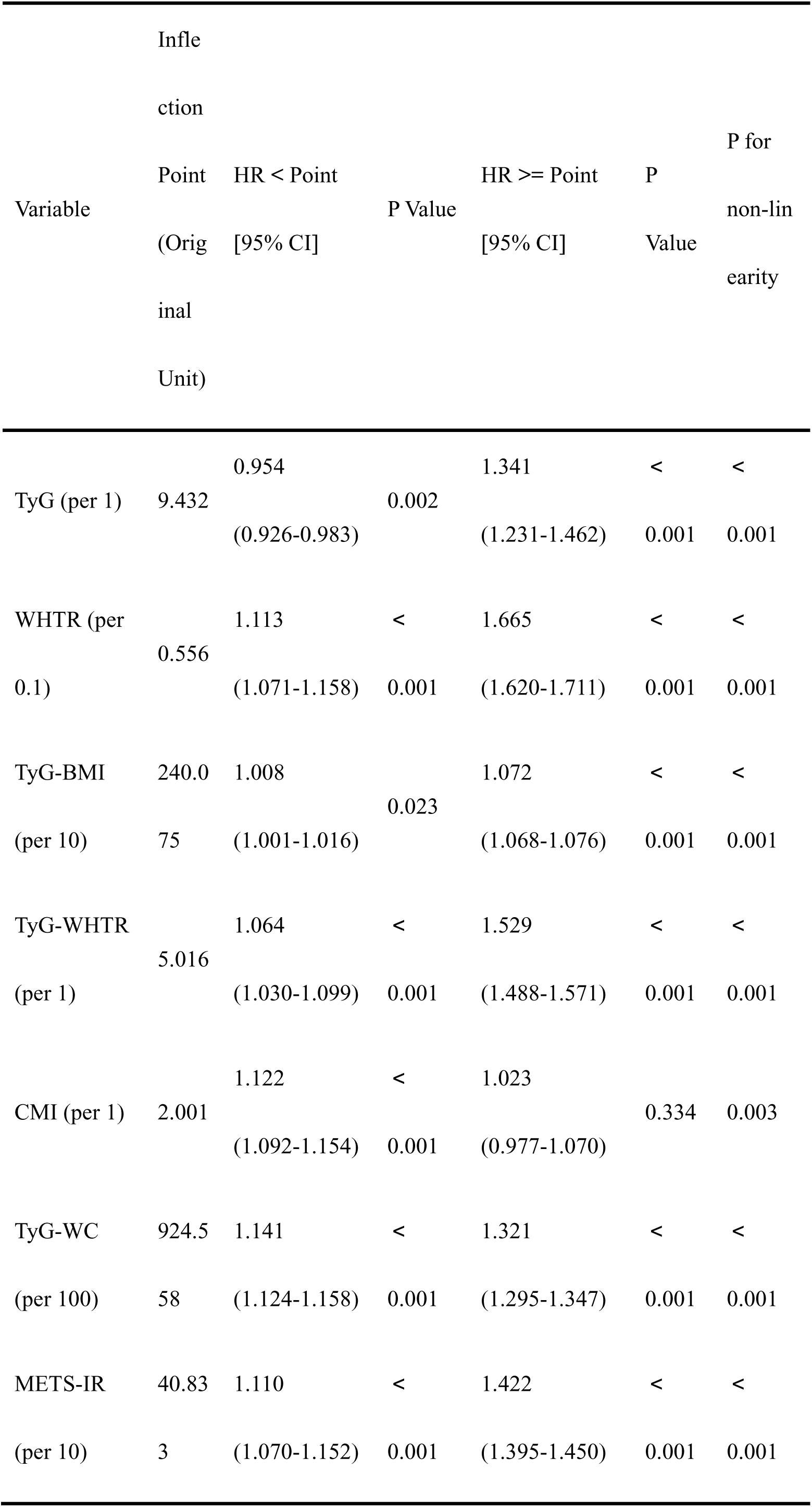
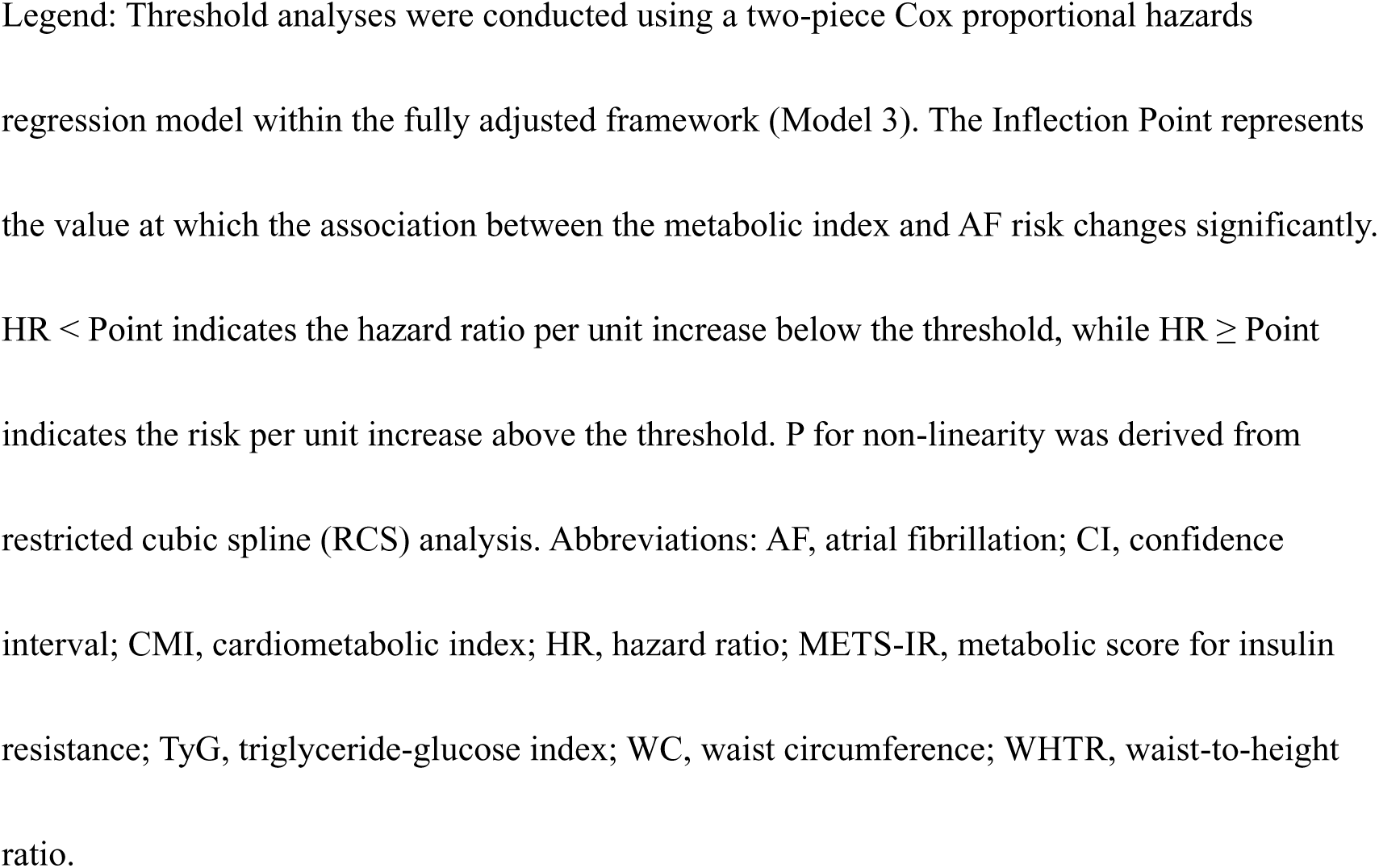
Threshold effect analysis of metabolic indices on atrial fibrillation risk using two-piece regression.

For WHTR, TyG-BMI, TyG-WHTR, TyG-WC, and METS-IR, we observed continuous non-linear positive associations, where the risk of AF escalated more rapidly after surpassing specific thresholds. The risk trajectory of WHTR steepened significantly beyond the inflection point of 0.556 (HR per 0.1-unit increase: 1.665 [1.620–1.711] above vs. 1.113 [1.071–1.158] below the threshold). Similarly, the hazard ratios were markedly higher above the inflection points. Specifically, for TyG-BMI (inflection point: 240.075), the risk increased slightly by 0.8% per 10-unit increment below the threshold but accelerated to 7.2% per 10-unit increment above it. A rapid accumulation of risk was also observed for TyG-WC (inflection point: 924.558) and METS-IR (inflection point: 40.833). Above their respective thresholds, the HRs were 1.321 per 100-unit increase for TyG-WC and 1.422 per 10-unit increase for METS-IR. These findings suggest that the pathogenic impact of insulin resistance combined with obesity on AF accumulates progressively but accelerates once metabolic burden exceeds a critical "tipping point." In contrast, the CMI index displayed a potential saturation effect. The risk of AF increased linearly up to an inflection point of 2.001 (HR: 1.122 per 1-unit increase, 95% CI: 1.092–1.154, P < 0.001). However, beyond this value, the association plateaued and lost statistical significance (HR: 1.023 per 1-unit increase, 95% CI: 0.977–1.070, P = 0.334). The detailed results from threshold analyses, including all inflection points, hazard ratios with 95% confidence intervals, and P-values for each segment, are provided in Table 3.

### 3.5 Predictive Value of Indices for Incident AF

To further evaluate and compare the predictive performance of the seven metabolic indices for the risk of incident AF, time-dependent ROC curve analyses were conducted at 3-, 5-, and 10-year follow-up intervals (Figure 4). Overall, the anthropometry-combined insulin resistance indices demonstrated superior discriminative ability compared to TyG or WHTR alone across all time points. Specifically, TyG-WC and METS-IR consistently exhibited the largest areas under the curve (AUCs), indicating a stronger predictive value for long-term AF risk.

**Figure 4.**
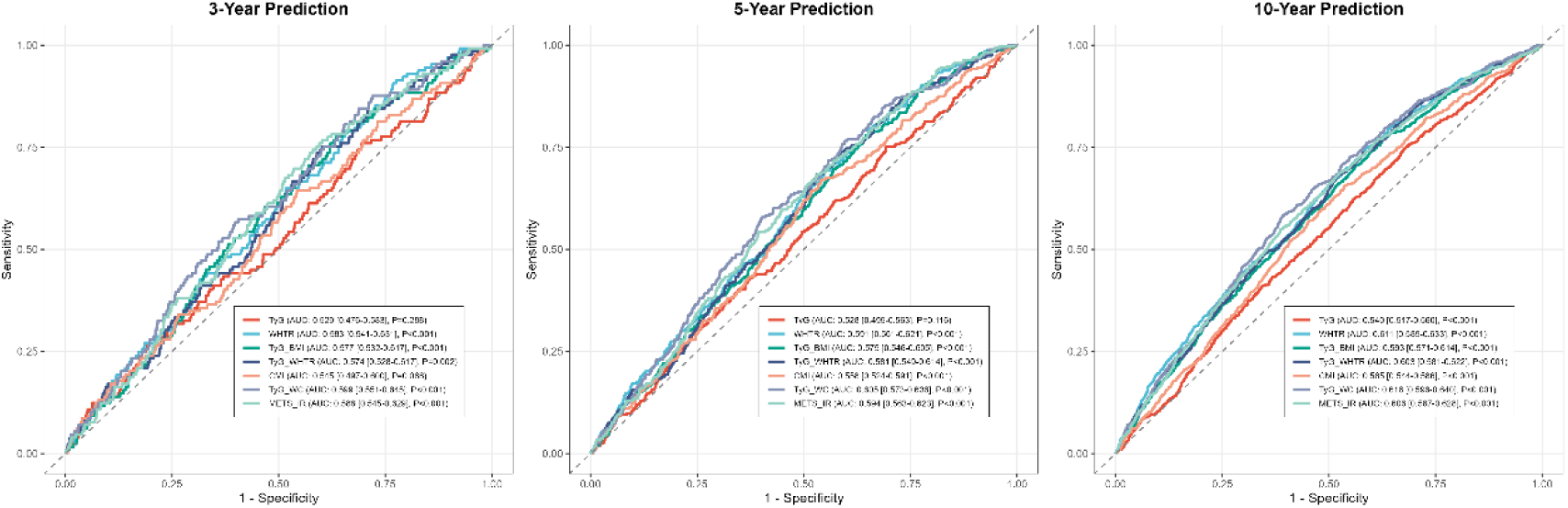
Time-dependent ROC curves comparing the predictive performance of metabolic indices for atrial fibrillation Receiver operating characteristic (ROC) curves illustrating the discriminatory ability of the TyG index versus anthropometry-combined indices for predicting incident AF at (A) 3 years, (B) 5 years, and (C) 10 years of follow-up. The Area Under the Curve (AUC) and 95% confidence intervals are provided for each index. Composite indices, particularly TyG-WC (dark blue) and METS-IR (teal), consistently demonstrated superior predictive accuracy (higher AUCs) compared to the standalone TyG index (red) or WHTR (light blue) across all time points. This superiority was statistically significant, highlighting the added value of integrating central adiposity measures.

In the 3-year prediction model, TyG-WC (AUC: 0.589; 95% CI: 0.554–0.624) and METS-IR (AUC: 0.588; 95% CI: 0.545–0.629) showed the highest predictive performance. Interestingly, the predictive ability of the TyG index alone was not statistically significant in the short-to-medium term (3-year AUC: 0.529, P = 0.200; 5-year AUC: 0.528, P = 0.116). In the 10-year prediction model, the discriminatory power of all indices improved or remained stable, with all associations reaching statistical significance (all P < 0.001). The composite indices maintained their dominance, with TyG-WC (AUC: 0.610; 95% CI: 0.588–0.632) and METS-IR (AUC: 0.608; 95% CI: 0.587–0.628) again outperforming the single TyG index (AUC: 0.549; 95% CI: 0.517–0.565). Collectively, these findings suggest that incorporating anthropometric parameters (such as WC and WHTR) into insulin resistance assessment significantly enhances the predictive capability for AF onset compared to using lipid-glucose parameters alone.

To further quantify the improvement in predictive accuracy conferred by incorporating anthropometric parameters into insulin resistance assessment, we calculated the continuous Net Reclassification Improvement (NRI) and Integrated Discrimination Improvement (IDI). Adding BMI or WC to the TyG index significantly improved risk stratification for incident AF. Compared to the TyG index alone, the TyG-BMI index yielded the most substantial improvement, with a continuous NRI of 0.118 (95% CI: 0.087-0.144, P < 0.001) and an IDI of 0.454% (95% CI: 0.280-0.695, P < 0.001) (Supplementary Table S2). This suggests that approximately 11.8% of participants were correctly reclassified into more appropriate risk categories when using TyG-BMI instead of TyG. Similarly, METS-IR and TyG-WHTR also demonstrated significant incremental predictive values (all P < 0.001 for NRI and IDI), confirming that the superior performance of these composite indices is not solely driven by statistical sensitivity but reflects a genuine enhancement in identifying high-risk individuals.

### 3.6 Interaction between genetic susceptibility and insulin resistance indices

To further explore whether the association between insulin resistance indices and AF risk is modified by genetic background, we performed stratified analyses based on PRS tertiles (Low, Intermediate, and High risk). The results of the interaction analyses are presented in Table 4 and Figure 5.

**Figure 5.**
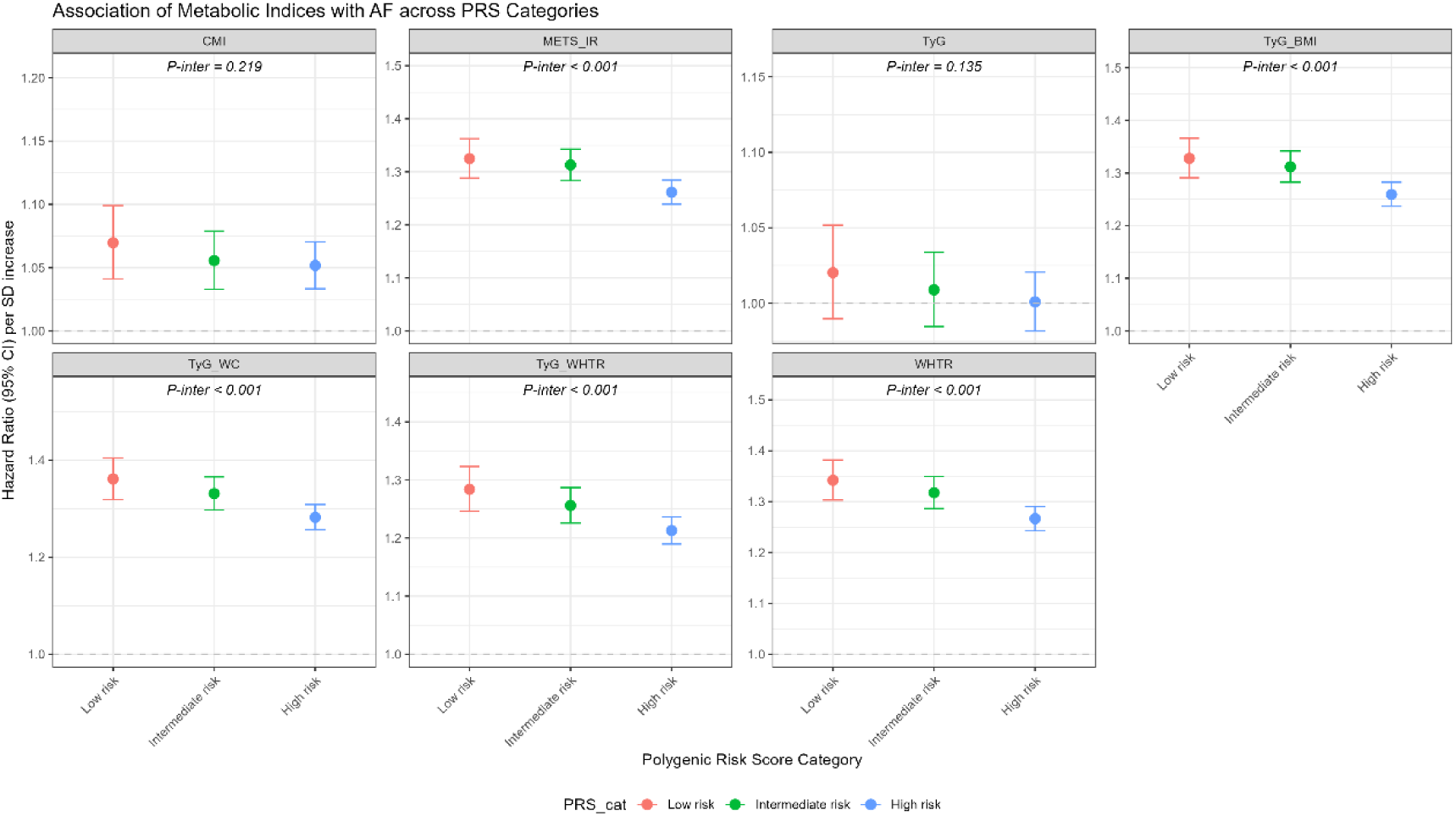
Stratified analyses by genetic susceptibility to atrial fibrillation Stratified analysis of the association between metabolic indices and AF risk across tertiles of the Polygenic Risk Score (PRS): Low (red), Intermediate (green), and High (blue) genetic risk. Data points represent the Hazard Ratio (HR) per 1-SD increase in each index, and error bars indicate 95% confidence intervals. P-inter values denote the statistical significance of the interaction between the metabolic index and PRS categories. Significant interactions (e.g., for TyG-WC, METS-IR) indicate that the relative risk conferred by metabolic dysfunction is significantly stronger in individuals with low genetic susceptibility compared to those with high genetic risk. The TyG index alone showed no significant interaction.

**Table 4.**
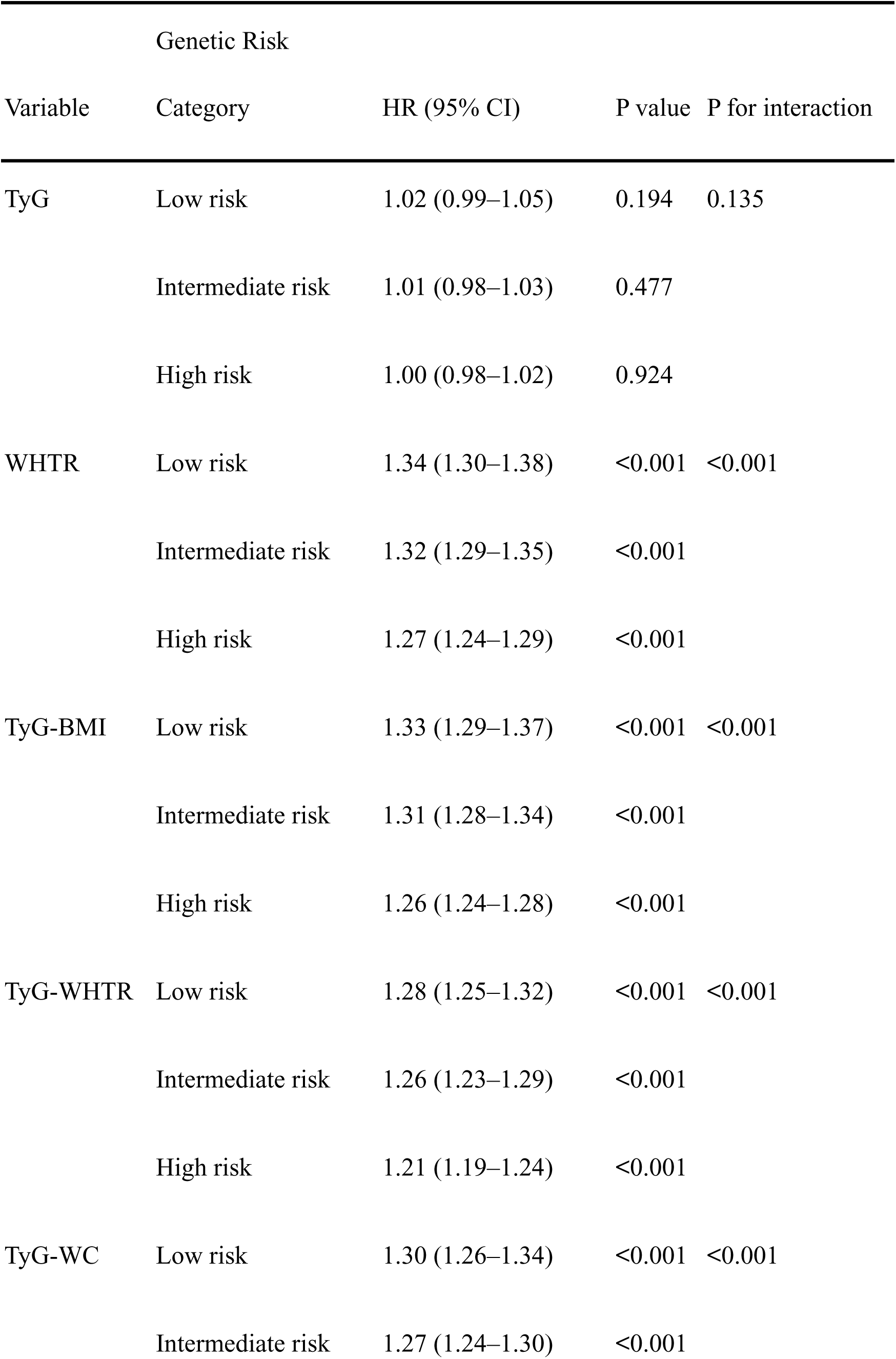
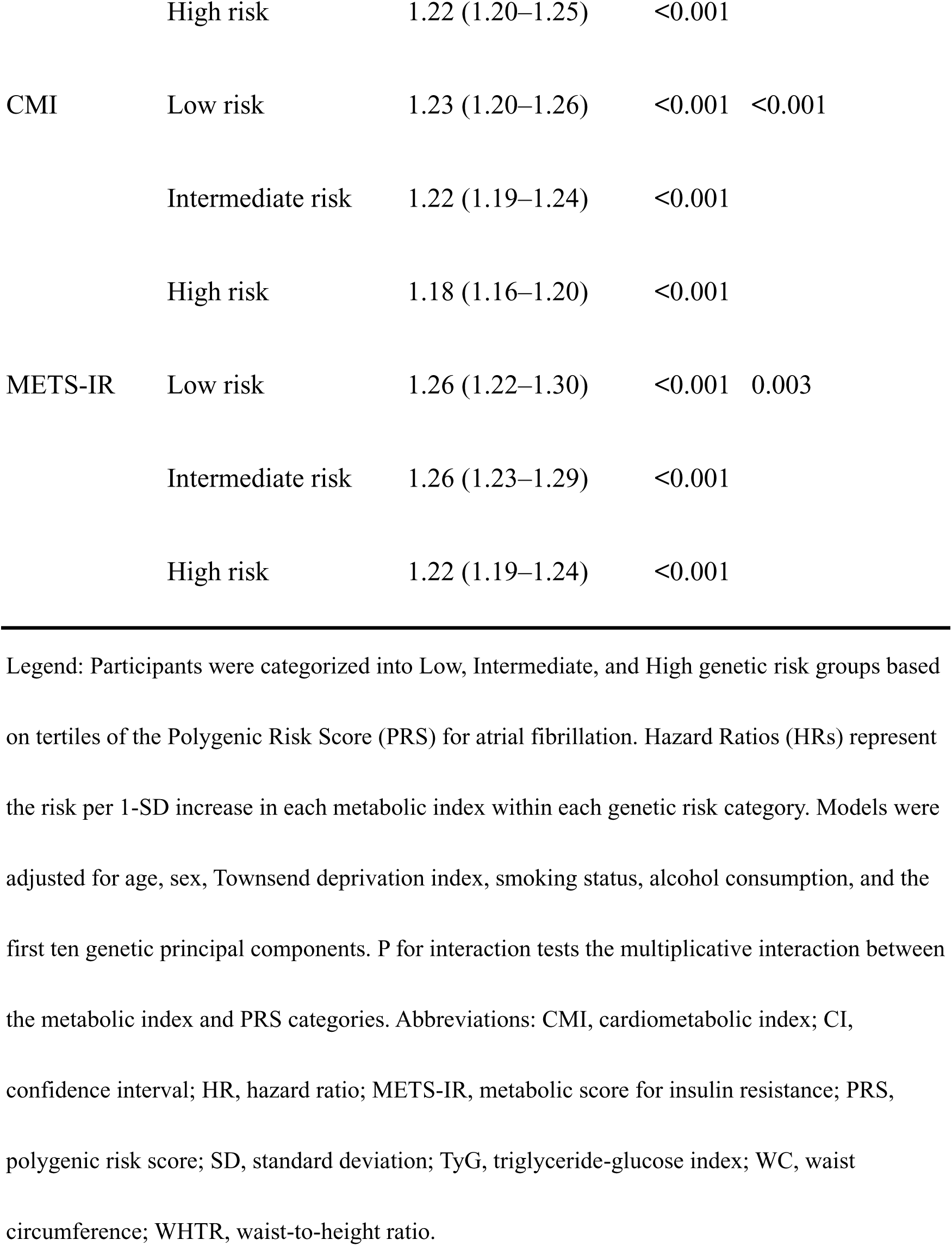
Stratified analyses of metabolic indices and atrial fibrillation risk by polygenic risk score tertiles.

**Table 5.** Associations of Metabolic Indices with Incident AF (Competing Risk Models) Hazard Ratios represent risk per 1-SD increase in exposure.

|  | **Model 1** |  |  | **Model 2** |  |  | **Model 3** |  |  |
| --- | --- | --- | --- | --- | --- | --- | --- | --- | --- |
| **Model 1**<br>**(Unadjusted)** | **HR** | **95% CI** | **p-value** | **HR** | **95% CI** | **p-value** | **HR** | **95% CI** | **p-value** |
| TyG (per SD) | 1.15 | 1.14,<br>1.17 | <0.001 | 1.01 | 1.00,<br>1.03 | 0.041 | 1.00 | 0.99,<br>1.02 | 0.6 |
| WHTR (per SD) | 1.44 | 1.43,<br>1.46 | <0.001 | 1.31 | 1.29,<br>1.32 | <0.001 | 1.29 | 1.27,<br>1.30 | <0.001 |
| TyG_BMI (per SD) | 1.33 | 1.32,<br>1.34 | <0.001 | 1.30 | 1.28,<br>1.32 | <0.001 | 1.28 | 1.27,<br>1.30 | <0.001 |
| TyG_WHTR (per SD) | 1.41 | 1.39,<br>1.42 | <0.001 | 1.25 | 1.23,<br>1.27 | <0.001 | 1.23 | 1.21,<br>1.25 | <0.001 |
| CMI (per SD) | 1.15 | 1.14,<br>1.16 | <0.001 | 1.07 | 1.05,<br>1.08 | <0.001 | 1.05 | 1.04,<br>1.06 | <0.001 |
| TyG_WC (per SD) | 1.48 | 1.47,<br>1.50 | <0.001 | 1.32 | 1.31,<br>1.34 | <0.001 | 1.30 | 1.28,<br>1.32 | <0.001 |
| METS_IR (per SD) | 1.34 | 1.33,<br>1.36 | <0.001 | 1.30 | 1.29,<br>1.32 | <0.001 | 1.28 | 1.27,<br>1.30 | <0.001 |

Abbreviations: CI = Confidence Interval, HR = Hazard Ratio

We observed significant interactions between genetic risk and anthropometry-based insulin resistance indices, including WHTR, TyG-BMI, TyG-WHTR, TyG-WC, and CMI (P for interaction < 0.001 for all), as well as METS-IR (P for interaction = 0.003). While the risk of AF significantly increased with elevated levels of these indices across all PRS categories, the magnitude of the association was notably more pronounced in participants with low genetic risk compared to those with high genetic risk, as shown in Figure 5. For instance, per 1-SD increase in TyG-BMI, the hazard ratio for AF was 1.33 (95% CI: 1.29–1.37) in the low PRS group, whereas it was slightly attenuated to 1.26 (95% CI: 1.24–1.28) in the high PRS group. A similar pattern was observed for TyG-WHTR (Low PRS: HR 1.28 vs. High PRS: HR 1.21) and TyG-WC (Low PRS: HR 1.30 vs. High PRS: HR 1.22). In contrast, the TyG index alone did not show a significant interaction with genetic susceptibility (P for interaction = 0.135), suggesting that the incorporation of obesity-related anthropometric parameters captures a risk pathway that is more distinct from, or interactive with, polygenic susceptibility.

### 3.7 Subgroup Analyses

To assess the consistency of the associations between IR indices and AF risk, we performed subgroup analyses stratified by age, sex, BMI, smoking status, hypertension, and diabetes mellitus. Figure 6A illustrates the results for TyG-WC, which demonstrated the most prominent predictive value among all evaluated parameters. The positive association between TyG-WC and the risk of new-onset AF remained statistically significant and generally consistent across all of strata. Notably, a significant interaction was observed between age group and TyG-WC (P for interaction < 0.001). The risk estimates were more pronounced in participants aged < 65 years [HR: 1.42(95% CI: 1.40-1.44) vs. HR: 1.24(95% CI: 1.21-1.27)] compared to the elderly population, implying that younger individuals might be more sensitive to the cumulative burden of insulin resistance and central obesity. In contrast, the traditional single TyG index failed to show a statistically significant association with AF risk in this population. As presented in Figure 6B, the confidence intervals for the TyG index crossed the null line across the majority of subgroups, and the overall predictive value was not significant (Overall HR: 1.01(95% CI: 0.99-1.02). This striking discrepancy highlights the critical role of abdominal obesity in AF pathogenesis. We also conducted parallel subgroup analyses for other metabolic parameters (WHTR, TyG-BMI, TyG-WHTR, CMI, and METS-IR), with detailed results presented in Supplementary Figures S1–S5. Specifically, incorporating central obesity metrics (BMI or WHTR) into the TyG index significantly enhanced the risk stratification capability, as evidenced by the higher hazard ratios and more robust confidence intervals observed in the TyG- BMI and TyG-WHTR groups compared to TyG alone. This finding reinforces the superiority of assessing insulin resistance and abdominal adiposity simultaneously. Our results suggest that insulin resistance alone (assessed by TyG) may not be a sufficient predictor for AF in this cohort, and incorporating anthropometric markers (WC or WHTR) is essential to unmask the true metabolic risk.

**Figure 6.**
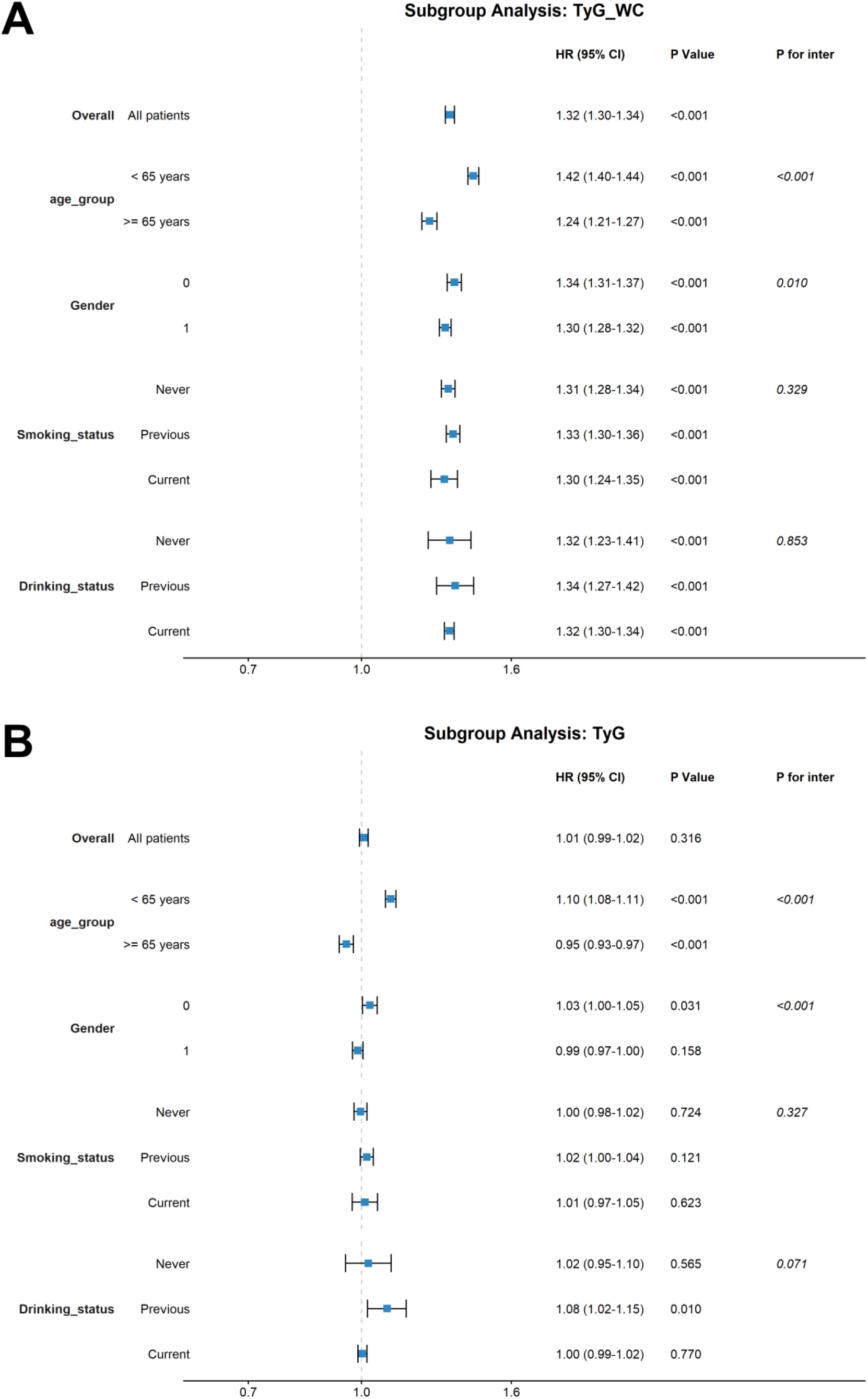
Subgroup analysis of the association between TyG-WC, TyG and the risk of new-onset atrial fibrillation. Forest plots displaying the multivariable-adjusted Hazard Ratios (HRs) for AF per 1-SD increase in (A) TyG-WC and (B) TyG index, stratified by key demographic and clinical factors (age, gender, smoking, drinking status). The blue squares represent point estimates, and horizontal lines indicate 95% confidence intervals. P-value indicates the significance of the association within each subgroup, while P for inter evaluates the heterogeneity of effects between subgroups. TyG-WC demonstrates robust and consistent positive associations across all strata. In contrast, the TyG index shows null associations in most subgroups (crossing the reference line of 1.0) and inconsistent patterns, further confirming the "TyG paradox."

### 3.8 Sensitivity and Robustness Analyses

We conducted a series of sensitivity analyses to verify the robustness of our primary findings. Firstly, to account for the potential competing risk of death (which may preclude the occurrence of AF), we employed the Fine-Gray subdistribution hazard model. As shown in Supplementary Figure S6, the results remained consistent with the primary Cox proportional hazards analysis. All the anthropometry-combined IR indices continued to show significant associations with AF risk (e.g., TyG-WC, TyG-WHTR, and METS-IR), maintaining higher subdistribution hazard ratios (SHRs) compared to single indices, confirming that the observed risks were not driven by survivor bias. Secondly, to minimize the potential bias of reverse causality (where latent AF might influence metabolic profiles before diagnosis), we conducted a lag analysis by excluding participants who developed AF within the first 2 and 3 years of follow-up (Supplementary Table S3). The hazard ratios remained virtually unchanged compared to the original analysis (e.g., TyG-WC HR: 1.32 after a 2-year lag and HR: 1.33 after a 3-year lag vs. original HR: 1.32). Thirdly, we calculated the E-value to assess the potential impact of unmeasured confounding (Supplementary Table S4). The E-value quantifies the minimum strength of association that an unmeasured confounder would need to have with both the exposure and the outcome to explain away the observed effect. The E-values for the association between all the anthropometry-combined IR indices (e.g., TyG-WC, METS-IR, and TyG-BMI) and AF were > 1.90. These values indicate that an unmeasured confounder would need to have a strong association (risk ratio > 1.90) with both the exposure and the outcome to explain away the observed results, suggesting the results are moderately robust to unmeasured confounding.

## 4. Discussion

Our large-scale prospective cohort study, leveraging data from the UK Biobank, provides compelling evidence that composite insulin resistance indices integrating measures of central adiposity, such as TyG-WC and METS-IR, offer superior and independent predictive value for incident atrial fibrillation compared to the singular lipid-glucose metric, the TyG index. The central finding, which we term the “TyG paradox,” reveals that while TyG showed an initial association with AF in unadjusted models, this relationship was substantially attenuated upon comprehensive multivariable adjustment. In stark contrast, indices such as TyG-WC, TyG-BMI, and METS-IR demonstrated robust, independent, and positive associations with AF risk. This observation underscores a critical nuance in metabolic risk assessment for AF: the biochemical signal of IR, as captured by TyG, may be insufficient on its own and requires contextualization within the framework of body composition, particularly visceral adiposity. The significant NRI of approximately 10-12% achieved by adding BMI or WC to TyG quantitatively affirms the necessity of a dual assessment strategy for AF risk stratification.

The superiority of composite indices aligns with the evolving pathophysiological understanding of AF as a condition deeply rooted in metabolic dysregulation and systemic inflammation^8,13,20,21^. Visceral adipose tissue is not a passive energy store but an active endocrine organ secreting pro-inflammatory adipokines and free fatty acids, which promote insulin resistance, endothelial dysfunction, and a pro-fibrotic state within the atrial myocardium^22^. Epicardial adipose tissue, a specific visceral fat depot, has been directly implicated in AF pathogenesis through paracrine effects^23,24^. Therefore, indices like TyG-WC or METS-IR likely capture this synergistic pathology more holistically than TyG alone, which reflects only the systemic insulin-resistant state. Our findings resonate with studies showing that metabolic syndrome and its components are potent drivers of AF^25–28^. For instance, the severe insulin-resistant diabetes (SIRD) subgroup exhibits a particularly high risk of AF^6^, and the burden of cardio-kidney-metabolic (CKM) domains strongly predicts adverse outcomes in AF patients^29^. The composite indices used in our study may serve as practical, integrative surrogates for these complex, multi-system pathological states.

A key advancement of this study is the identification of clear nonlinear threshold effects for these composite indices. The restricted cubic spline analyses revealed distinct inflection points, such as 924.6 for TyG-WC and 40.8 for METS-IR, beyond which AF risk escalates sharply. This nonlinear relationship has significant clinical implications, suggesting the existence of a metabolic “tipping point.” Below this threshold, interventions might yield incremental benefits, but surpassing it could trigger a cascade of pathological remodeling leading to a steep rise in AF susceptibility. This concept moves beyond linear risk gradients and provides a potential quantitative target for intensive lifestyle or pharmacological intervention aimed at pushing an individual’s metabolic profile back below the high-risk threshold. Similar nonlinear associations have been observed for TyG and other indices in relation to cardiovascular outcomes, including in AF populations^12,30^, supporting the biological plausibility of such threshold phenomena. The mechanisms may involve exceeding a capacity for metabolic homeostasis, leading to accelerated oxidative stress, inflammation, and direct lipotoxic effects on cardiomyocytes^31,32^.

Perhaps the most novel aspect of our investigation is the exploration of genetic-environment interaction. We found that elevated composite metabolic indices conferred a significantly increased AF risk across all strata of polygenic risk, demonstrating that the detrimental effect of poor metabolic health is universal and not confined to those with high genetic predisposition. This underscores the broad applicability and potential impact of lifestyle interventions targeting obesity and insulin resistance for AF prevention, regardless of an individual’s genetic background^33^. More importantly, we detected significant multiplicative interactions between the PRS and composite indices like TyG-WC and TyG-BMI, an effect absent for the standalone TyG index. This interaction was characterized by a steeper increase in relative risk associated with metabolic dysfunction among individuals at low genetic risk. This pattern suggests that the composite indices are capturing a distinct, environmentally modifiable pathway to AF—specifically, the synergy of visceral adiposity and insulin resistance—that operates more independently of the polygenic background driving AF through other mechanisms (e.g., ion channelopathies, structural heart disease predisposition). In individuals with high genetic risk, AF may develop through these strong innate pathways regardless of metabolic status, whereas in those with low genetic risk, the “metabolic pathway” becomes the predominant driver. This aligns with the concept of heterogeneous pathways to AF^14,34^ and suggests that composite metabolic indices are particularly valuable for identifying individuals whose AF risk is primarily metabolically driven.

Our results must be interpreted in the context of the existing literature, which presents a complex picture regarding TyG and AF. ^8,13,35–37^. For example, a recent meta-analysis concluded that higher TyG is consistently associated with increased AF risk^8^, and TyG has been linked to left atrial appendage thrombogenic milieu^38^. However, other analyses, including one within the UK Biobank focusing on insulin sensitivity measured by estimated glucose disposal rate (eGDR), highlight the protective role of insulin sensitivity^39^. The apparent discrepancy with our finding of a null association for adjusted TyG may stem from differences in study population characteristics, follow-up duration, and crucially, the extent of covariate adjustment. Many prior studies adjusted for a limited set of confounders, whereas our fully adjusted models included detailed anthropometric measures. This suggests that the association of TyG with AF may be largely mediated or confounded by adiposity measures. When these are accounted for, the unique contribution of the TyG variable itself diminishes. This interpretation is supported by the superior performance of TyG-BMI in machine learning models predicting AF in comorbid populations^10,40^ and its prognostic value in patients with AF and metabolic syndrome^30^. Furthermore, our findings on the value of composite indices are corroborated by research on the metabolic score for visceral fat (METS-VF), which was also prospectively linked to AF risk in the UK Biobank^11^.

The clinical implications of this work are substantial. First, it argues for a shift in preventive cardiology towards using integrated metrics like TyG-WC or METS-IR in AF risk assessment, especially in primary care and cardiometabolic clinics. These indices are calculated from routine, low-cost measurements (waist circumference, BMI, fasting triglycerides, and glucose), making them highly scalable. Second, the identified thresholds offer concrete targets for patient counseling and goal-setting.

Third, the evidence of gene-environment interaction reinforces the message that metabolic health is a powerful lever for AF prevention across the entire genetic risk spectrum. This is particularly encouraging for public health initiatives, as it suggests that population-level strategies to combat obesity and insulin resistance could meaningfully reduce the burden of AF. The protective effect of improved insulin sensitivity, as suggested by eGDR^33^, and the benefits of exercise in mitigating metabolic atrial remodeling^31^ provide a mechanistic basis for such interventions. The integration of such simple indices into digital health platforms or digital twin models could further personalize risk prediction and management^41^.

This study has several strengths, including its prospective design, large sample size, long follow-up, detailed phenotypic data, and the incorporation of genetic information. However, limitations must be acknowledged. Despite extensive adjustment, residual confounding from unmeasured or imperfectly measured factors (e.g., diet quality, physical activity intensity, sleep patterns) is possible. The UK Biobank population is not fully representative of the general population, being generally healthier and less ethnically diverse, which may affect the generalizability of the absolute risk estimates and specific threshold values. The PRS used captures common genetic variants but not rare variants or gene-gene interactions. Furthermore, while we demonstrate association and improved risk classification, further studies are needed to establish whether interventions that lower these composite indices directly lead to reduced AF incidence.

Future research should validate the proposed threshold values in independent, multi-ethnic cohorts. Mechanistic studies are warranted to elucidate the precise biological pathways linking high TyG-WC or METS-IR to atrial electrophysiological and structural remodeling, potentially focusing on adipokine profiles, mitochondrial function, and inflammatory cascades^4,42^. Investigating the dynamic changes in these indices over time and their relationship to AF risk would be highly informative. Additionally, interventional trials testing whether targeted reductions in these composite metrics—through weight loss, improved diet, or specific pharmacotherapies—can prevent AF, especially in individuals above the identified risk thresholds, would be the ultimate test of their clinical utility. The field of predictive, preventive, and personalized medicine stands to benefit greatly from such integrative, pathophysiology-informed biomarkers^40,41^.

In conclusion, our findings advocate for a paradigm shift in assessing metabolic risk for atrial fibrillation. The isolated triglyceride-glucose index, while a useful marker of insulin resistance, provides an incomplete picture. Composite indices that marry the biochemical signal of insulin resistance with the anatomical reality of central obesity, such as TyG-waist circumference, offer superior prediction, reveal critical risk thresholds, and uniquely capture a modifiable pathway that interacts with genetic predisposition. These readily available tools can enhance early identification of high-risk individuals and strengthen the evidence base for targeted, lifestyle-centered prevention strategies aimed at curbing the growing global epidemic of atrial fibrillation.

## Conclusions

Our study establishes that composite metrics integrating central adiposity (specifically TyG-WC and TyG-BMI) provide superior AF risk prediction compared to the TyG index alone, effectively resolving the "TyG paradox." Beyond linear associations, we identified critical risk thresholds (e.g., TyG-WC > 924.6) that signal a sharp escalation in AF susceptibility. Importantly, these metabolic risks operate independently of polygenic susceptibility, suggesting that targeting metabolic health—through weight management and insulin sensitization—remains a primary prevention strategy, offering substantial benefits even in populations with low genetic risk.

## Data Availability

Data supporting the findings of this study from the UK Biobank team (http://www.ukbiobank.ac.uk/). The data and methods that support the findings of this study are available from the corresponding author on reasonable request.

## List of abbreviations

AF: Atrial fibrillation
AUC: Area under the curve
BMI: Body mass index
CI: Confidence interval
CKM: Cardiovascular-kidney-metabolic
CMI: Cardiometabolic index
CRP: C-reactive protein
DBP: Diastolic blood pressure
eGDR: Estimated glucose disposal rate
GWAS: Genome-wide association study
HbA1c: Glycated hemoglobin
HR: Hazard ratio
ICD-10: International Classification of Diseases, 10th Revision
IDI: Integrated discrimination improvement
IR: Insulin resistance
LDL-C: Low-density lipoprotein cholesterol
METS-IR: Metabolic score for insulin resistance
METS-VF: Metabolic score for visceral fat
NRI: Net reclassification improvement
PRS: Polygenic risk score
RCS: Restricted cubic splines
SBP: Systolic blood pressure
SD: Standard deviation
SIRD: Severe insulin-resistant diabetes
TDI: Townsend deprivation index
TyG: Triglyceride-glucose index
TyG-BMI: Triglyceride-glucose-body mass index
TyG-WC: Triglyceride-glucose-waist circumference
TyG-WHTR: Triglyceride-glucose-waist-to-height ratio
WBC: White blood cell
WC: Waist circumference
WHTR: Waist-to-height ratio

## Declarations

### Ethics approval and consent to participate

The UK Biobank was approved by the North West Multicenter Research Ethics Committee, with all participants providing written informed consent. Ethical approval and informed consent were waived as the UK Biobank data is publicly available and does not include identifiable information.

### Consent for publication

Not applicable

### Authors’ contributions

Z.K. and W.SW. performed the data analyses and wrote the manuscript. Z.K., SF.Z. and W.S. prepared all the tables and figures. W.SW., H.M., and R.CW. helped perform the analysis with constructive discussions. C.H. and L.YQ. contributed to the conception of the study. All authors read and approved the final manuscript.

## Acknowledgments

We thank all participants and staff from the UK Biobank study. Hao Cui served as the Principal Investigator of the UK Biobank project (Application ID: 545415) that granted access to the data used in this study.

## Authors’ information

Department of Cardiovascular Surgery, Beijing Anzhen Hospital, Capital Medical University

Beijing Anzhen Hospital, Capital Medical University, The Key Laboratory of Remodeling-Related Cardiovascular Diseases, Ministry of Education, Beijing Institute of Heart, Lung and Blood Vessel Diseases, Beijing 100029, China

Ke Zhang, Shengwei Wang, Song Wang, Shifeng Zhao, Meng He, Changwei Ren; Hao Cui, Yongqiang Lai

## Supplementary Information

Additional file 1

File format: .docx

Title of data: Supplementary Table S1

Description of data: Table presenting the detailed numbers of at-risk participants at each follow-up time point across quartiles of the seven metabolic and anthropometric indices.

Additional file 2

File format: .xlsx

Title of data: Supplementary Table S2

Description of data: Table showing the results of Net Reclassification Improvement (NRI) and Integrated Discrimination Improvement (IDI) analyses, quantifying the incremental predictive value of adding anthropometric metrics to the TyG index.

Additional file 3

File format: .xlsx

Title of data: Supplementary Table S3

Description of data: Table displaying sensitivity analyses (lag analyses) excluding participants diagnosed with atrial fibrillation within the first 2 and 3 years of follow-up to assess potential reverse causality.

Additional file 4

File format: .xlsx

Title of data: Supplementary Table S4

Description of data: Table presenting E-value calculations to assess the potential impact of unmeasured confounding on the observed associations between metabolic indices and atrial fibrillation.

Additional file 5

File format: .pdf

Title of data: Supplementary Figures S1–S5

Description of data: A collection of supplementary figures containing subgroup analyses for all metabolic indices (Figs. S1–S5).

Additional file 6

File format: .tiff

Title of data: Supplementary Figure S6

Description of data: Forest plot showing the sensitivity analysis using the Fine-Gray subdistribution hazard model to account for the competing risk of death.

